# Immunomodulation by intravenous omega-3 fatty acid treatment in older subjects hospitalized for COVID-19: a single-blind randomized controlled trial

**DOI:** 10.1101/2021.12.27.21268264

**Authors:** Hildur Arnardottir, Sven-Christian Pawelzik, Philip Sarajlic, Alessandro Quaranta, Johan Kolmert, Dorota Religa, Craig E. Wheelock, Magnus Bäck

**Author notes:** Shared first author (Hildur Arnardottir, Sven-Christian Pawelzik). **Registration in Trial Registries**: This trial “Resolving Inflammatory Storm in COVID-19 Patients by Omega-3 Polyunsaturated Fatty Acids - A single-blind, randomized, placebo-controlled feasibility study” (COVID-Omega-F) is registered in the European Union Drug Regulating Authorities Clinical Trials (EudraCT) database with number 2020-002293-28 and at Clinical Trials.gov with number NCT04647604. **Data availability**: Individual participant data that underlie the results reported will be shared, after deidentification, with researchers who provide a methodologically sound proposal. *Time Frame*: Beginning 9 months following article publication and finishing 36 months following article publication. *Access Criteria*: Investigators interested in data should contact the corresponding author. **Author for Correspondence** Professor Magnus Bäck, Karolinska University Hospital, M85, 14186 Stockholm, Sweden.

## Abstract

Severe acute respiratory syndrome coronavirus 2 (SARS-CoV-2) causes coronavirus disease 2019 (COVID-19) with respiratory distress and systemic hyperinflammation. The primary objective of this single-blind randomized controlled proof-of-concept clinical trial was to establish the effects of intravenous (i.v.) omega-3 (n-3) polyunsaturated fatty acid (PUFA) treatment compared to placebo on inflammatory markers in COVID-19, represented by leukocytes as well as inflammatory protein and lipid mediators. Here we also present an exploratory analysis of the mechanisms of action to elucidate the potential to resolve the COVID-19 hyperinflammation through interfering with lipid mediators. Inclusion criteria were COVID-19 diagnosis and clinical status requiring hospitalization. Randomization was 1:1 to a once daily i.v. infusion (2 mL/kg) of either n-3 PUFA emulsion containing 10g of fish oil per 100 mL or placebo (NaCl) for 5 days. Results from 22 older subjects (mean age 81±6.1 years) were analyzed. The neutrophil to lymphocyte ratio was significantly decreased after n-3 PUFA administration. Liquid chromatography–mass spectrometry (LC-MS/MS) -based lipid metabolite analysis established increased proresolving lipid mediator precursor levels and decreased formation of leukotoxin and isoleukotoxin diols by n-3 PUFA treatment. The mechanistic exploration revealed decreased immunothrombosis and preserved interferon-response. Finally, n-3 PUFA treatment may serve to limit cortisone-induced immunosuppression, including preserving leukocyte phagocytic capacity. In conclusion, i.v. n-3 PUFA administration was safe and feasible during hospitalization of multimorbid older subjects for COVID-19. The results identified n-3 PUFA treatment mediated lipid signature of increased proresolving precursor levels and decreased leukotoxin diols in parallel to beneficial immune responses. EudraCT: 2020-002293-28; clinicaltrials.gov: <u>NCT04647604</u>.

## Introduction

The uncontrolled inflammatory response in coronavirus disease 2019 (COVID-19) is characterized by a high neutrophil to lymphocyte ratio (NLR) and increased circulating levels of inflammatory mediators, represented by a monocytic cytokine release syndrome (1) and increased proinflammatory lipid mediators (2). Systemic uncontrolled inflammation is common in severe SARS-CoV2 infection and a potential therapeutic target to improve prognosis in COVID-19 (3, 4). Older subjects represent a particularly vulnerable patient population in COVID-19. The introduction of anti-inflammatory cortisone treatment (5) has confirmed the benefit of immunomodulation, although with the possible caveat of also causing immunosuppression (6). In addition to inhibiting proinflammatory signaling, acute inflammation could potentially be disrupted by a functional and effective resolution to limit and eventually turn off inflammation while retaining an intact host defense.

Fatty acids are crucial in controlling inflammation. A disturbed balance in endogenous PUFA metabolism increases proinflammatory and leukotoxic lipid mediators with concomitant decrease in proresolving lipid mediators (7). The omega-3 (n-3) polyunstaurated fatty acids (PUFA) eicosapentaenoic acid (EPA) and docosahexaenoic acid (DHA) decrease proinflammatory lipid mediators and serve as precursors for lipid mediators of the resolution of inflammation that dampen systemic inflammation while also improving healing and microbial elimination (7). The recent VASCEPA-COVID-19 CardioLink-9 trial showed that oral n-3 PUFA treatment with the EPA ethyl ester icosapent ethyl for 14 days improves symptoms and reduces C-reactive protein (CRP) in ambulatory COVID-19 patients (8). Similar results have been reported in a smaller study of critically ill COVID-19 patients (9), however excluding patients not receiving enteral nutrition (10). Intravenous (i.v.) n-3 PUFA treatment reduces hyperinflammation in other critical infectious conditions (11) but has not previously been studied in COVID-19.

The primary objective of this randomized controlled proof-of-concept clinical trial was to establish the effects of i.v. n-3 PUFA compared to placebo on inflammatory markers in COVID-19, represented by leukocytes as well as inflammatory protein and lipid mediators (12). Here we also present an exploratory analysis of the mechanisms of action to elucidate the potential to resolve the inflammatory storm in COVID-19 through interfering with lipid mediators.

## Methods

### Patients

The trial “Resolving inflammatory storm in COVID-19 patients by Omega-3 Polyunsaturated fatty acids” (COVID-Omega-F) was approved by Swedish Ethical Review Authority on May 25, 2020 (Dnr 2020-02592) and by the Medical Product Agency on May 29, 2020 (Dnr 5.1-2020-42861). An amendment was approved to increase inclusion to achieve comparable groups completing the full study protocol according to the initial sample size calculations (Swedish Ethical Review Authority approval November 25, 2020; Dnr 2020-06137, and Medical Product Agency approval on November 30, 2020; Dnr 5.1-2020-96391). Inclusion criteria were COVID-19 diagnosis and clinical status requiring hospitalization. After signed informed consent, participants were randomized 1:1 to a once daily i.v. infusion (2 mL/kg) of either placebo (0.9% NaCl) or n-3 PUFA emulsion (Omegaven® bought from ApoEX, Stockholm, Sweden) containing 10 g of fish oil per 100 mL, of which 1.25-2.82 g DHA and 1.44-3.09 g EPA for 5 days. Randomization was performed using sequentially numbered sealed envelopes in random permuted blocks of n=2-3. Participants were blinded to the intervention. The sample size was determined to at least 10 patients in each group based on previous studies of i.v. PUFA emulsions on infectious inflammation (11). The primary endpoint measures were changes in inflammatory biomarkers (time frame: after either 5 days treatment or at study end, whichever came first) defined as () white blood cell counts, () C-reactive protein (CRP), () lipidomic profiling, and () cytokines. The study protocol registered at clinicaltrials.gov with reference NCT04647604 has been published (12) and is provided in the Appendix of the present report.

### Blood and urine sample collection

Laboratory measures of routine biochemistry, blood cell counts, and CRP were performed by the Karolinska University Laboratory in accordance with ISO15189. Blood and urine samples for biomarker and lipid metabolite analyses were collected at the study start before administration of the first dose of treatment (baseline), at 24-48h after the first administered dose (early), and within 24h of the last administered dose (end). Plasma was used for measurements of markers of inflammation using the multiplex Ella(tm) system (ProteinSimple, Biotechne, San Jose, CA, USA).

### Lipid metabolite analysis

Lipid mediator quantification was performed as previously described (13) with minor modifications to the sample preparation to account for the solvent composition. One mL MeOH was added to 250 µL plasma and samples were vortexed and centrifuged. The supernatant was transferred to glass tubes and evaporated under gentle N_2_ stream to remove the MeOH before reconstitution in 1 mL of extraction solution (0.2 M Na_2_HPO_4_ / 0.1 M citric acid, pH 5.6) followed by solid phase extraction as previously reported (13). Twelve additional compounds were included in the current study that were not reported in the previous methods description, including epoxides and vicinal diols from AA and DHA (Supplementary Table 1). Non-commercially epoxide and vicinal diol analytical standards were kindly provided by Bruce Hammock at the University of California Davis (CA, USA) and synthesized by Dr Johanna Revol-Cavalier at Karolinska Institutet.

### Blood cell isolation

Whole blood was collected into an 8 mL sodium heparinized CPT vacutainer and processed within 2 h of collection. Peripheral blood mononuclear cells (PBMC) were separated from erythrocytes and neutrophils following centrifugation at 1800 x g for 15 min at room temperature. Erythrocytes were isolated by 3% dextran sedimentation for 20 minutes at room temperature and then washed 3 times with PBS^-/-^ (followed each time by centrifugation at 1000 x g, 10 min, 4°C).

### PBMC stimulations for mediator release measurements

PBMC were collected as described above, washed with PBS^-/-^ followed by centrifugation for 10 min at 250 x g, 4°C, and then resuspended in RPMI media containing 1% heat-inactivated (56°C for 30 min) autologous serum at a concentration of 2.5×10^6^ cells/mL. 0.5×10^6^ cells/well were seeded into a round bottom 96-well plate and incubated with or without 1 µg/mL LPS (serotype O111:B4, Sigma-Aldrich, Merck KGaA, St. Louis, MO, USA) for 6 h. Following centrifugation for 5 min at 300 x g, 4°C, the supernatants were collected and stored at -80°C.

### Luminex

Supernatants from stimulated and unstimulated PBMC were used for measurements of markers of inflammation using Human XL Cytokine Luminex Performance Assay multiplex kit (RnD Systems, Minneapollis, MO, USA).

### Flow cytometric analysis of stimulated whole blood

Fresh whole blood was obtained from sodium heparinized CPT vacutainers prior to centrifugation for whole blood functional assays. For phagocytosis assessment, 30 µl whole blood was incubated with opsonized pH sensitive zymosan particles (pHrodo(tm) Red, Invitrogen, Thermo Fisher Scientific, Waltham, MA, USA) for 60 min at 37°C or 4°C (negative control). Samples were then incubated with APC-conjugated anti-human CD66b antibody, FITC-conjugated anti-human CD14 antibody, and eFluor450-conjugated anti-human CD45 antibody (all from Thermo Fisher Scientific, Waltham, MA, USA) for 15 min on ice. For platelet-leukocyte aggregation assessment, 45 µl whole blood were incubated with 10 µM ADP (Sigma-Aldrich, Merck KGaA, St. Louis, MO, USA) for 10 minutes at 37°C. Samples were then incubated with APC-conjugated anti-human CD66b antibody, PE-conjugated anti-human CD41a antibody, FITC-conjugated anti-human CD14 antibody, and eFluor450-conjugated anti-human CD16 antibody (all from Thermo Fisher Scientific, Waltham, MA, USA) for 15 min at RT. All samples were fixed and lysed using 1-step Fix/Lyse solution (eBioscience, Thermo Fisher Scientific, Waltham, MA, USA) and washed 2 time with Flow Cytometry Staining Buffer (eBioscience, from Thermo Fisher Scientific, Waltham, MA, USA). Cells were analyzed with BD Fortessa flow cytometer (BD Biosciences) and Flow Jo software version v10.7.0.

### Measurement of biological interferon activity

An interferon (IFN) sensitive human amniotic epithelial cell line (WISH, ATCC® CCL-25(tm)) was used to assess the biological activity of type I (IFN-α and IFN-β) and type II (IFN-γ) IFN, respectively, in patient serum as previously described (14). In brief, 1.25×10^5^ cells/well in a total volume of 200 µl MEM supplemented with 5% heat-inactivated FBS, 2 mM L-glutamine, 1 mM Na-pyruvate, 10 mM HEPES, 50 IU/ml penicillin, and 50 µg/ml streptomycin (all from Thermo Fisher Scientific, Waltham, MA, USA), were seeded into a 96-well plate and cultured over night at 37°C in a humified 5% CO_2_ atmosphere. At the day of the experiment, the medium was aspirated and replaced with 200 µl MEM supplemented with 50% patient sera. Control cells were incubated with serum-free medium containing 0-200 IU/ml recombinant human IFN-α, IFN-β, and IFN-γ, respectively (RnD Systems, Minneapolis, MO, USA). After incubation for 6 h at 37°C in a humified 5% CO_2_ atmosphere, the media were aspirated, the cells were lyzed in 350 µl RLT buffer (Qiagen, Hilden, Germany) supplemented with 10 µl/ml β -mercaptoethanol and frozen at -80°C until further analysis. Total RNA was extracted using the RNeasy Mini Kit (Qiagen, Hilden, Germany) following the manufacturer’s instruction and subsequently reverse transcribed into cDNA using the High-Capacity RNA-to-cDNA Kit (Thermo Fisher Scientific, Waltham, MA, USA). Expression levels of the type I IFN-induced response gene MX1 and the type II IFN-induced response gene CXCL9 were analyzed by quantitative real time PCR (qPCR) on a QuantStudio 7 Flex Real-Time PCR System using specific TaqMan Gene expression assays (MX1, Hs00895608_m1; CXCL9, Hs00171065_m1, all from Thermo Fisher Scientific, Waltham, MA, USA) and quantified by comparison to the expression levels in the respective standard curves obtained from cells that were incubated with recombinant IFN protein. The procedure was replicated with WISH cells that were cultured for three consecutive passages prior to the experiment in MEM supplemented with 0.01% n-3 PUFA emulsion (v/v) to enrich n-3 PUFA in the cell membrane.

### Statistical analysis

Outcome measures are expressed as means with standard errors. Key results are presented as separate line plots for the placebo and n-3 PUFA groups showing changes within subjects between baseline and the endpoint. Paired t-tests were used to determine significance in changes over time. To determine the effect of time and the n-3 PUFA intervention, a mixed model analysis of variance was implemented in which time was defined as a within-subjects effect and the intervention as a between-subjects effect. Moreover, the interaction between these effects was measured. Due to the limited statistical power of the study, descriptive outcomes were not tested for significance as defined in the prespecified analysis plan in the study protocol, which is provided as an Appendix online. To gain insight into the key lipid metabolites separating the placebo and n-3 PUFA intervention groups, we implemented an orthogonal partial least squares discriminant analysis (OPLS-DA) model to analyze the variation correlated to the intervention. The residual variation is projected upon the components that are orthogonal to the main component that is predictive of the target of interest. The model generated variable statistic (R2) and approximation (Q2) importance scores to identify what metabolites accounted for the greatest differences in the placebo and n-3 PUFA groups. All significance tests were two-sided and findings with P<0.05 were regarded as statistically significant. Statistical analyses were performed in R version 4.1.1 (R Core Team 2021). R: A language and environment for statistical computing. R Foundation for Statistical Computing, Vienna, Austria. URL https://www.R-project.org/), Bioconductor version 3.13 (15), GraphPad (version 8.4.3, Sand Diego, California, USA), and SigmaPlot for Windows Version 14.5.

## Results

### Patient characteristics

All subjects were included between from June to December 2020, before COVID-19 vaccination was available, at a first COVID-19 infection. The flow chart of patient inclusion is shown in Supplementary Fig 1. The study was completed for 22 older subjects (mean age 81±6.1 years). There were no significant differences in baseline characteristics between the groups (Table 1). The number of administered doses were on average 4.3±1.5 in the intervention group (n=10) and 4.5±1.0 in the placebo group (n=12; P=0.66). Seven patients in each group received concomitant corticoids (P=0.68). There were 2 in-hospital deaths in each group. The n-3 PUFA treatment was administered in the form of triglycerides, and as a safety measure, fasting serum triglyceride levels were monitored daily during the study and not significantly increased. No serious adverse events were encountered.

**Table 1.** Patient characteristics. Demographic data are expressed as either mean ± standard deviation or numbers and per cent. Laboratory measures are expressed as median (interquartile range). Statistical analyses of demographic data were performed by either a Student’s t-test (continuous variables) or a Fisher Exact Test (categorical variables). Cardiovascular disease was defined as medical history of at least one of the following: coronary artery disease, cerebrovascular disease, peripheral artery disease, heart failure, atrial fibrillation, venous thromboembolism. All laboratory measures were carried out by Karolinska University Laboratory in accordance with ISO15189. Statistical analyses of laboratory data were performed using a Mann-Whitney Rank Sum Test. *<u>Abbreviations:</u> ALAT, alanine aminotransaminase; APTT, activated partial thromboplastin time; ASAT, aspartate aminotransferase; COPD, chronic obstructive pulmonary disease; CRP, C-reactive protein; DOAC, direct oral anticoagulants, eGFR, estimated glomerular filtration rate; EVF, erythrocyte volume fraction, ICS, inhaled corticosteroids, IL, interleukin; LD, lactate dehydrogenase; NLR, neutrophil-to-lymphocyte ratio; RAS, renin-angiotensin system*.

|  | *All* | *Placebo* | *n-3 PUFA* | *P* |
| --- | --- | --- | --- | --- |
| *n* | 22 | 12 | 10 |  |
| *Age - years* | 81.1±6.1 | 81.5±5.6 | 80.7±7.0 | 0.77 |
| *Female sex – no. (%)* | 12 (55%) | 7 (58%) | 5 (50%) | 0.70 |
| *Body Mass Index - kg/m<sup>2</sup>* | 25.6±3.2 | 25.0±5.6 | 26.3±4.4 | 0.46 |
| *Current smoker – no. (%)* | 2 (9%) | 0 (0%) | 2 (20%) | 0.20 |
| *Days since symptom start* | 7.1±3.3 | 7.2±3.5 | 7.0±3.1 | 0.87 |
| *Days since COVID-19 diagnosis* | 3.5±1.7 | 3.5±2.0 | 3.7±1.3 | 0.83 |
| ***Medical history – no. (%)*** |  |  |  |  |
| *Cardiovascular disease* | 14 (64%) | 9 (75%) | 5 (50%) | 0.38 |
| *Hypertension* | 15 (64%) | 9 (75%) | 6 (60%) | 0.65 |
| *COPD/Asthma* | 3 (13%) | 1 (8.3%) | 2 (20%) | 0.57 |
| *Cancer* | 4 (18%) | 2 (17%) | 2 (20%) | 1.0 |
| *Diabetes* | 9 (41%) | 3 (24%) | 6 (60%) | 0.19 |
| *CKD* | 5 (23%) | 2 (17%) | 3 (30%) | 0.62 |
| *Rheumatological disease* | 4 (18%) | 2 (17%) | 2 (20%) | 1.0 |
| ***Concomitant medication – no. (%)*** |  |  |  |  |
| *Acetyl salicylic acid* | 8 (36%) | 6 (50%) | 2 (20%) | 0.20 |
| *Clopidogrel* | 3 (14%) | 2 (17%) | 1 (10%) | 1.0 |
| *Agents acting on the RAS* | 9 (41%) | 5 (42%) | 4 (40%) | 1.0 |
| *Beta blockers* | 11 (50%) | 5 (42%) | 6 (60%) | 0.67 |
| *Calcium channel blockers* | 3 (14%) | 2 (17%) | 1 (10%) | 1.0 |
| *Diuretics* | 5 (23%) | 3 (26%) | 2 (20%) | 1.0 |
| *Nitrates* | 2 (9%) | 0 (0%) | 2 (20%) | 0.20 |
| *Statins* | 10 (45%) | 5 (42%) | 5 (50%) | 1.0 |
| *DOAC* | 5 (23%) | 1 (8%) | 4 (40%) | 0.14 |
| *Insulin* | 4 (18%) | 2 (17%) | 2 (20%) | 1.0 |
| *Oral anti-diabetics* | 5 (23%) | 1 (8%) | 4 (40%) | 0.14 |
| *ICS* | 3 (14%) | 1 (8%) | 2 (20%) | 0.57 |
| ***Blood Cell Counts*** |  |  |  |  |
| *Leukocytes (x10<sup>9</sup>/L)* | 6.7 (4.7-9.6) | 5.6 (3.7-9.2) | 8.1 (6.1-11) | 0.09 |
| *Monocytes (x10<sup>9</sup>/L)* | 0.6 (0.3-0.8) | 0.45 (0.30-0.78) | 0.65 (0.50-0.85) | 0.26 |
| *Neutrophils (x10<sup>9</sup>/L)* | 5.0 (3.2-6.7) | 4.0 (2.3-6.3) | 5.6 (4.1-9.2) | 0.06 |
| *Lymphocytes (x10<sup>9</sup>/L)* | 1.1 (0.8-1.5) | 1.0 (0.85-1.48) | 1.15 (0.78-1.62) | 0.72 |
| *NLR* | 3.7 (2.5-8.1) | 3.3 (2.1-7.3) | 4.7 (3.2-11.5) | 0.28 |
| *Erythrocytes (x10<sup>12</sup>/L)* | 4.1 (3.4-4.5) | 4.2 (3.4-4.7) | 4.0 (3.4-4.3) | 0.36 |

**Table 1. Patient characteristics.**
|  |  |  |  |  |
| --- | --- | --- | --- | --- |
| *EVF* | 0.36 (0.32-0.40) | 0.35 (0.30-0.40) | 0.36 (0.34-0.39) | 0.70 |
| *Platelets (x10<sup>9</sup>/L)* | 226 (168-292) | 217 (162-306) | 240 (190-289) | 0.90 |
| **<u>Inflammation, Infection</u>** |  |  |  |  |
| *CRP (mg/L)* | 64 (37-87) | 65 (40-92) | 62 (20-75) | 0.53 |
| *IL-6 (ng/L)* | 34 (11-53) | 47 (26-54) | 14 (2.5-72) | 0.09 |
| *Procalcitonin (μg/L)* | 0.13 (0.08-0.27) | 0.10 (0.07-0.18) | 0.13 (0.12-0.57) | 0.57 |
| **<u>Coagulation, Thrombosis</u>** |  |  |  |  |
| *Fibrin-D-dimer (mg/L)* | 0.71 (0.57-1.78) | 0.61 (0.51-0.92) | 1.1 (0.67-1.9) | 0.08 |
| *Fibrinogen (g/L)* | 5 (3.9-5.6) | 5.0 (4.1-5.3) | 4.8 (3.5-6.2) | 0.92 |
| *APTT (s)* | 26 (23-29) | 24 (23-26) | 25 (22-34) | 0.65 |
| *INR* | 1.1 (1.0-1.2) | 1.1 (1.0-1.1) | 1.1 (1.1-1.4) | 0.33 |
| **<u>Organ Damage</u>** |  |  |  |  |
| *ALAT (μcat/L)* | 0.34 (0.27-0.76) | 0.33 (0.23-0.68) | 0.36 (0.28-1.31) | 0.45 |
| *ASAT (μcat/L)* | 0.43 (0.36-0.83) | 0.45 (0.37-0.83) | 0.43 (0.28-0.89) | 0.95 |
| *LD (μcat/L)* | 5.0 (3.8-6.2) | 5.0 (4.0-6.9) | 4.8 (3.6-5.8) | 0.55 |
| *Bilirubin (μmol/L)* | 7 (6-12) | 6 (5-10) | 9.5 (6-16) | 0.09 |
| *Creatinine (μmol/L)* | 72 (59-106) | 80 (59-95) | 65 (55-114) | 0.90 |
| *eGFR (mL/min/1.73 m<sup>2</sup>)* | 59 (45-77) | 59 (47-74) | 61 (45-81) | 0.87 |
| *Troponin T (ng/L)* | 19 (13-39) | 15 (13-47) | 24 (13-33) | 0.92 |
| **<u>Chemistry</u>** |  |  |  |  |
| *Hb (g/L)* | 116 (102-129) | 111 (96-132) | 120 (106-129) | 0.60 |
| *Ferritin (μmol/L)* | 614 (223-810) | 674 (213-799) | 498 (232-1578) | 0.88 |
| *Na (mmol/L)* | 138 (136-140) | 139.5 (136- 140) | 137 (134-140) | 0.50 |
| *K (μmol/L)* | 3.9 (3.7-4.3) | 3.8 (3.7-4.1) | 4.2 (3.8-4.5) | 0.20 |
| *Ca (μmol/L)* | 2.21 (2.12-2.29) | 2.21 (2.12-2.29) | 2.21 (2.12-2.29) | 0.80 |
| *Albumin (g/L)* | 28 (26-32) | 30 (27-33) | 28 (24-30) | 0.26 |

### Primary endpoint measures

#### White blood cell counts

The neutrophil to lymphocyte ratio (NLR) was significantly decreased after n-3 PUFA administration (Fig 1), while it was not significantly altered in the placebo group. The mean increase in lymphocyte count was 0.4±0.08×10^9^/L in the intervention group and 0.03±0.16×10^9^/L in the placebo group (P=0.03). There were no significant changes in total leukocyte, neutrophil or monocyte counts (data not shown).

**Fig 1.**
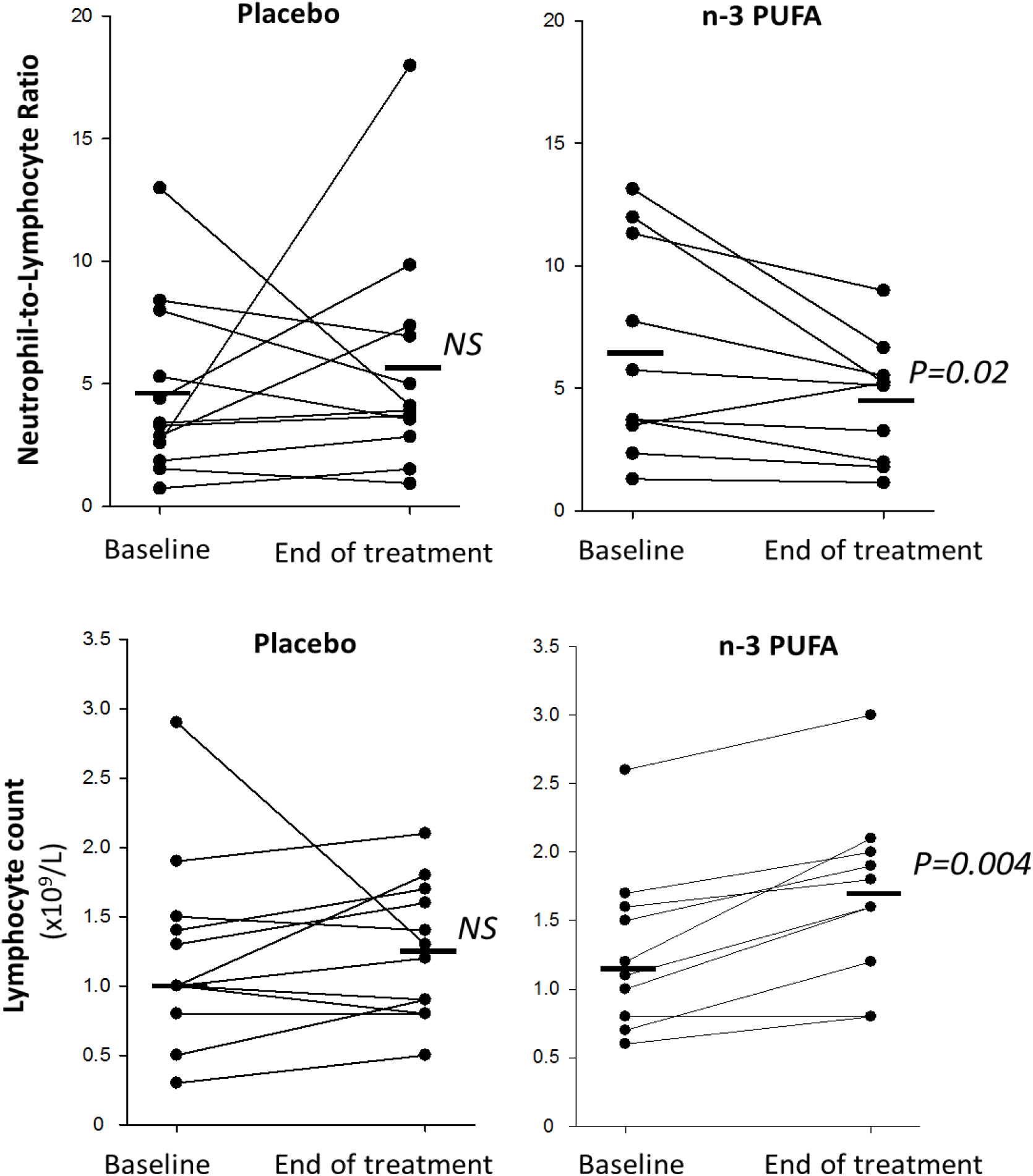
Neutrophil-to-lymphocyte ratio (upper panels) and lymphocyte counts (lower panels) at baseline and after treatment (End) with intravenous infusion (2 mL/kg) of either placebo (NaCl; left panels, n=12) or n-3 PUFA emulsion containing 10 g of fish oil per 100 mL (right panels, n=10). Horizontal lines represent the median at each time point. Statistical analyses were performed with a Wilcoxon Signed Rank Test for repeated measures. NS, non-significant. Statistical significance was set at P=0.05.

#### C-reactive protein

CRP decreased from 62 (20-75) to 19 (3.8-27) mg/L after n-3 PUFA (P=0.08) and from 65 (40-92) to 32 (24-40) mg/L after placebo treatment (P=0.08).

#### Increased plasma n-3 PUFA concentrations

Patients randomized to n-3 PUFA treatment exhibited significant increase in plasma concentrations of EPA and DHA compared with the placebo group (Fig 2A). At the end of the study, EPA- and DHA concentrations were increased 3.7 (2.5-5.4) fold and 2.0 (1.5-2.5) fold from baseline, respectively in these patients. In contrast, the EPA and DHA plasma levels were not significantly altered in the placebo group; EPA 1.0 (0.8-1.2) and DHA 1.0 (0.9-1.2) -fold at the end of the study as compared to baseline (Fig 2A). The plasma concentrations of the n-6 PUFA arachidonic acid (AA) were not significantly altered from baseline and not significantly different between the n-3 PUFA and placebo group (Fig 2A).

**Fig 2.**
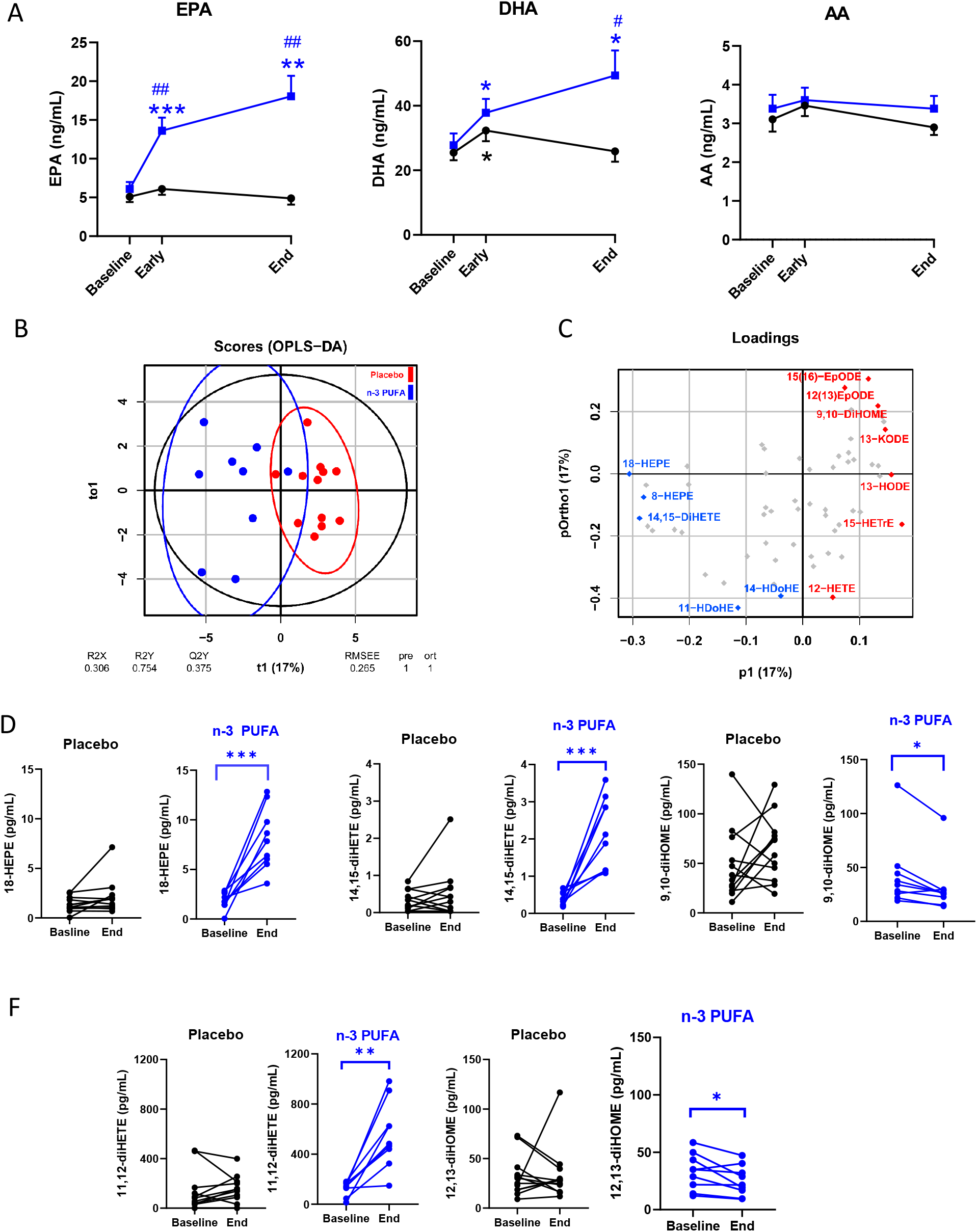
**(A)** Mass spectrometry analysis of circulating free fatty acids, eicosapentaenoic acid (EPA), docosahexanoic acid (DHA), and arachidonic acid (AA) in plasma from patients at baseline, 48 h (Early) and after treatment (End) following intravenous infusion (2 mL/kg) of either placebo (NaCl; black symbols, n=12) or n-3 PUFA emulsion containing 10 g of fish oil per 100 mL (blue symbols, n=10) for 5 days. Results are expressed as mean ± SEM. Statistical analyses were performed with 2-way ANOVA for repeated measures and post hoc testing. **(B)** Orthogonal partial least squares discriminant analysis (OPLS-DA) separated the n-3 PUFA (blue symbols) and placebo (red symbols). **(C)** The loading plot identified the EPA metabolites 18-HEPE (left), 14,15-diHETE (middle), and the LA metabolite 9,10-diHOME (right) among the critical molecules explaining the differences in plasma lipid mediator profile caused by n-3 PUFA intravenous infusion (blue= higher and red = lower in the n-3 PUFA group), which were **(D)** significantly altered in the n-PUFA (blue symbols; n=9) but not placebo (black symbols; n=12) groups between baseline and End. (F) Change in the corresponding EPA and LA diols, 11,12-diHETE (middle) and 12,13-diHOME (right). Results are presented as individual values and statistical analysis was performed by a paired Student’s t test. Statistical significance was set at P=0.05. * p<0.05; **p<0.01; ***p<0.001 compared to baseline and ^#^p<0.05; ^##^p<0.01 between placebo and n-3 PUFA treatment.

#### Altered plasma lipid metabolite profile by n-3 PUFA administration in COVID-19

The n-3 PUFA and placebo groups evidenced a separation in OPLS-DA based upon quantified levels of lipid metabolites, with a statistic fit (R2=0.75) higher that its approximation (Q2=0.38) to a testing dataset (Fig 2B). The DHA- and EPA-derived lipid metabolite profiles displayed in Table 2 revealed a significant increase in n-3 PUFA-derived metabolites, with EPA metabolites exhibiting the most pronounced increases in the n-3 PUFA compared with placebo groups (Table 2). The arachidonic acid (AA) -derived lipid metabolite profile is shown in Supplementary Table 2, and the octadecanoid pathways from linoleic acid (LA), α-LA, γ-LA, in Supplementary Table 3. The variable importance calculated from the OPLS-DA (Fig 2B) identified 18-HEPE, 14,15-diHETE, and 9,10-diHOME among the molecules explaining the differences in plasma lipid mediator profile between n-3 PUFA and placebo treatment (Fig 2C). These metabolites (Fig 2D) and the corresponding EPA and LA diols 11,12-diHETE and 12,13-diHOME (Fig 2F), were significantly altered after n-3 PUFA but not placebo treatment.

**Table 2.** Lipid mediator metabolites (pg/mL) from the n-3 PUFA EPA and DHA in patient plasma. P-values were derived from ta mixed effects ANOVA containing intervention (Interv; n-3 PUFA vs. placebo) and progress (early and end samples) as a within-subjects effect and as a between subjects effect. Intervention:progress (Interv:Prog) represents the significance of the interaction between these two terms *Abbreviations: PG, Prostaglandin; TX, Abbreviations: PG, Prostaglandin; TX, thromboxane; EPE, Hydroxyeicosapentaenoic acid; EpETE, Epoxyeicosatetraenoic acid. DiHETE, Dihydroxyeicosatetraenoic acid; HDoHE, Hydroxydocosahexaenoic acid; HDoHE, Hydroxydocosahexaenoic acid; EpDPA, Epoxydocosapentaenoic acid; DiHDPA, Dihydroxydocosapentaenoic acid*

|  | n-PUFA (n=9) |  |  |  | Placebo (n=12) |  |  | P-values |  |  |  |
| --- | --- | --- | --- | --- | --- | --- | --- | --- | --- | --- | --- |
|  | Baseline | Early | End |  | Baseline | Early | End |  | Interv | Progress | Interv:Progr |
| EPA Metabolites |  |  |  |  |  |  |  |  |  |  |  |
| PGE<sub>3</sub> | 0.08 (0.06) | 0.14 (0.08) | 0.36 (0.16) |  | 0.05 (0.03) | 0.11 (0.06) | 0.02 (0) |  | 0.064 | 0.394 | 0.051 |
| TXB<sub>3</sub> | 0.08 (0) | 0.83 (0.5) | 0.5 (0.42) |  | 0.08 (0) | 0.08 (0) | 0.08 (0) |  | 0.13 | 0.258 | 0.258 |
| 5-HEPE | 3.96 (0.77) | 6.28 (0.88) | 7.9 (1.39) |  | 3.3 (0.49) | 3.38 (0.41) | 3.08 (0.55) |  | 0.002 | 0.181 | 0.058 |
| 8-HEPE | 0.39 (0.06) | 1.1 (0.15) | 2.2 (0.32) |  | 0.32 (0.08) | 0.29 (0.06) | 0.51 (0.2) |  | 0.0000175 | 0.000639 | 0.014 |
| 9-HEPE | 0.2 (0.07) | 0.83 (0.27) | 1.97 (0.34) |  | 0.25 (0.08) | 0.19 (0.07) | 0.25 (0.13) |  | 0.0000727 | 0.003 | 0.005 |
| 11-HEPE | 0.26 (0.06) | 0.7 (0.11) | 1.44 (0.27) |  | 0.2 (0.07) | 0.2 (0.08) | 0.26 (0.09) |  | 0.0000927 | 0.002 | 0.005 |
| 12-HEPE | 20.89 (4.53) | 34.37 (6.03) | 60.9 (18.62) |  | 13.5 (2.71) | 17.36 (4.3) | 18.1 (4.59) |  | 0.007 | 0.117 | 0.138 |
| 15-HEPE | 5.05 (1.63) | 7.16 (1.69) | 7.17 (1.11) |  | 1.72 (0.57) | 2.96 (0.82) | 3.1 (0.96) |  | 0.015 | 0.87 | 0.892 |
| 18-HEPE | 1.86 (0.29) | 5.33 (0.59) | 8.11 (1.04) |  | 1.38 (0.22) | 1.9 (0.17) | 2.12 (0.49) |  | 0.0000044 | 0.001 | 0.005 |
| 8(9)-EpETE | 12.88 (12.86) | 5.76 (5.74) | 26.1 (26.08) |  | 5.09 (5.07) | 25.4 (18.9) | 0.02 (0) |  | 0.839 | 0.877 | 0.171 |
| 11(12)-EpETE | 88.07 (45.09) | 208.73 (79.04) | 329.04 (77.2) |  | 143. (56.4) | 83.1 (63.5) | 94.8 (40.5) |  | 0.033 | 0.175 | 0.26 |
| 14(15)-EpETE | 0.05 (0.02) | 0.26 (0.13) | 0.27 (0.08) |  | 0.12 (0.05) | 0.04 (0) | 0.04 (0) |  | 0.014 | 0.937 | 0.937 |
| 17(18)-EpETE | 0.22 (0.15) | 1.35 (0.39) | 1.7 (0.44) |  | 0.2 (0.13) | 0.13 (0.11) | 0.32 (0.21) |  | 0.003 | 0.073 | 0.584 |
| 5,6-DiHETE | 0.16 (0.06) | 0.08 (0) | 0.09 (0.01) |  | 0.08 (0) | 0.08 (0) | 0.08 (0) |  | 0.11 | 0.258 | 0.258 |
| 11,12-DiHETE | 125.46 (19.19) | 439.42 (80.77) | 555.99 (88.35) |  | 129 (47.1) | 102 (34.8) | 145 (33.1) |  | 0.0000965 | 0.023 | 0.267 |
| 14,15-DiHETE | 5.05 (0.42) | 9.54 (1.54) | 13.1 (1.76) |  | 4.73 (0.88) | 3.9 (0.78) | 4.16 (0.77) |  | 0.0000733 | 0.037 | 0.069 |
| 17,18-DiHETE | 39.95 (4.84) | 84.24 (14.08) | 116.99 (20.56) |  | 40.0 (4.12) | 41.3 (3.7) | 41.3 (4.66) |  | 0.000842 | 0.006 | 0.006 |
| DHA Metabolites |  |  |  |  |  |  |  |  |  |  |  |
| 8-HDoHE | 3.38 (0.53) | 4.09 (0.76) | 6.28 (1.03) |  | 2.41 (0.23) | 3.03 (0.3) | 2.99 (0.33) |  | 0.011 | 0.013 | 0.01 |
| 11-HDoHE | 5.03 (0.82) | 4.94 (0.89) | 8.21 (1.97) |  | 2.94 (0.63) | 4.36 (0.97) | 4.38 (0.93) |  | 0.124 | 0.126 | 0.132 |
| 14-HDoHE | 62.54 (11.02) | 61.9 (14.97) | 96.86 (28.46) |  | 48.0 (7.94) | 68.7 (17.0) | 69.4 (16.4) |  | 0.652 | 0.273 | 0.292 |
| 17-HDoHE | 36.82 (6.91) | 43.17 (5.51) | 41.93 (4.14) |  | 23.3 (3.29) | 33.6 (4.82) | 36.0 (5.7) |  | 0.267 | 0.843 | 0.545 |
| 7(8)-EpDPA | 99.62 (58.02) | 161.95 (81.96) | 152.14 (79.64) |  | 85.6 (36.4) | 39.6 (24.5) | 79.5 (36.5) |  | 0.19 | 0.635 | 0.436 |
| 10(11)-EpDPA | 1604.71 (237.47) | 2142.73 (373.51) | 2201.72 (318.09) |  | 1810 (294) | 1817 (240) | 1774 (312) |  | 0.392 | 0.93 | 0.603 |
| 16(17)-EpDPA | 74.56 (53.14) | 133.47 (56.71) | 191.08 (59.28) |  | 30.5 (24.7) | 58.9 (26.02) | 30.8 (23.3) |  | 0.035 | 0.564 | 0.104 |
| 19(20)-EpDPA | 1.84 (0.67) | 4.4 (1.55) | 5.75 (1.57) |  | 2.52 (0.54) | 2.95 (0.5) | 3.3 (0.75) |  | 0.196 | 0.09 | 0.301 |

**Table 2. Lipid mediator metabolites (pg/mL) from the n-3 PUFA EPA and DHA in patient plasma.**
|  | n-PUFA (n=9) |  |  |  | Placebo (n=12) |  |  |  | P-values |  |  |
| --- | --- | --- | --- | --- | --- | --- | --- | --- | --- | --- | --- |
| *7,8-DiHDPA* | 114.17 (58.64) | 144.49 (80.12) | 354.16 (140.23) |  | 65.4 (44.6) | 99.3 (52.2) | 132 (59.3) |  | 0.134 | 0.15 | 0.285 |
| *10,11-DiHDPA* | 18.62 (18.6) | 33.73 (27.84) | 48.07 (25.37) |  | 9.03 (9.01) | 6.99 (5.82) | 3.79 (2.08) |  | 0.131 | 0.401 | 0.192 |
| *16,17-DiHDPA* | 18.61 (10.97) | 78.29 (35.56) | 96.12 (35.85) |  | 76.5 (24.0) | 72.4 (26.8) | 54.7 (17.3) |  | 0.529 | 0.998 | 0.286 |
| *19,20-DiHDPA* | 12.77 (2.07) | 18.29 (2.57) | 24.48 (4.81) |  | 16.15 (1.6) | 16.71 (1.6) | 15.7 (1.16) |  | 0.127 | 0.153 | 0.05 |
| *16,17-DiHDPA* | 18.61 (10.97) | 78.29 (35.56) | 96.12 (35.85) |  | 76.5 (24.0) | 72.4 (26.8) | 54.7 (17.3) |  | 0.529 | 0.998 | 0.286 |
| *10,11-DiHDPA* | 18.62 (18.6) | 33.73 (27.84) | 48.07 (25.37) |  | 9.03 (9.01) | 6.99 (5.82) | 3.79 (2.08) |  | 0.131 | 0.401 | 0.192 |

#### No significant changes in plasma cytokines

The levels of the plasma cytokines interleukin (IL)-1β, IL-6, and TNF-α were not significantly altered in either of the groups during the study duration (Supplementary Fig 2).

### Exploratory outcome measures for mechanistic pathways

#### Reduced platelet leukocyte interactions

Immunothrombosis has been implicated as a key pathophysiological consequence of SARS-CoV-2 infection (16). To evaluate the n-3 PUFA effects on immunothrombosis in the present trial, *ex vivo* ADP-stimulation of whole blood was performed followed by flow cytometry analysis for platelet-leukocyte aggregates. Blood from n-3 PUFA-treated COVID-19 patients displayed a significantly reduced platelet-leukocyte aggregates at end of treatment from baseline compared with blood from the placebo group (Fig 3A). This change in platelet-leukocyte aggregates was driven by a significant decrease in the formation of platelet-neutrophil aggregates in the n-3 PUFA group that was not observed in the placebo group (Fig 3B-C). Since the platelet-leukocyte aggregation is mediated through monocyte-derived platelet activators, we next investigated the release of thrombogenic mediators from PBMC isolated from all trial participants. A significantly lower release of the immuno-thrombosis mediator Platelet-Derived Growth Factor (PDGF) and Regulated on Activation, Normal T Expressed and Secrete (RANTES) release, was observed in LPS-stimulated PBMC isolated from the n-3 PUFA group compared with the placebo group (Fig 3D).

**Fig 3.**
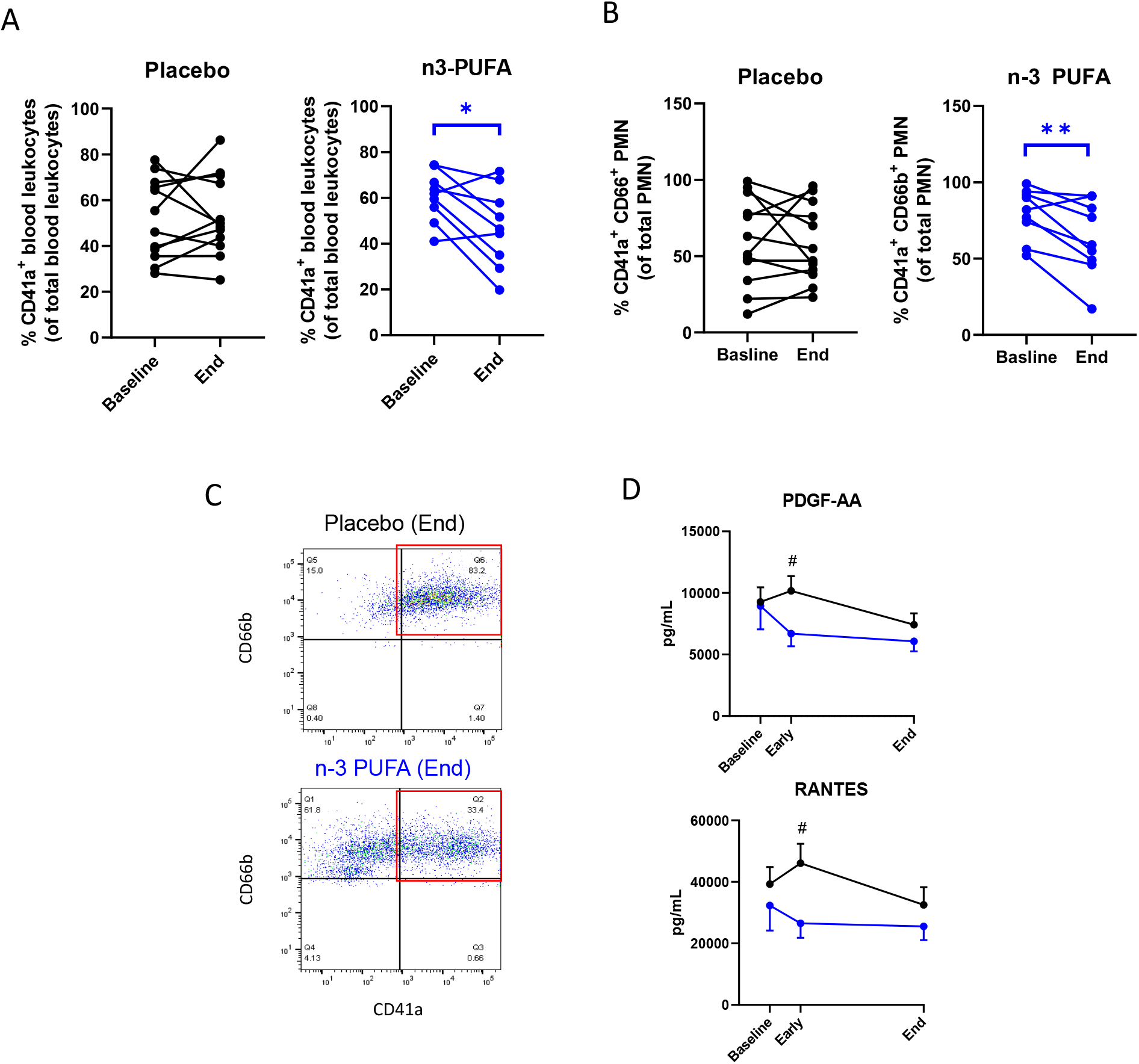
**(A)** Platelet-leukocyte aggregates and **(B)** Platelet-neutrophil aggregates at baseline and after treatment (End) with intravenous infusion (2 mL/kg) of either placebo (NaCl; black symbol, n=12) or n-3 PUFA emulsion containing 10 g of fish oil per 100 mL (blue symbol, n=9). Results are presented as individual values and statistical analysis was performed by a paired Student’s t test. (C) Representative flow cytometry dot blots showing CD41+ CD66b+ platelet-neutrophil aggregates in blood from Placebo (top) and n-3 PUFA (bottom) treated patients. (D) The release of PBMC-derived platelet activators, platelet derived growth factor (PDGF)-AA and regulated on activation, normal T cell expressed and secreted, RANTES from LPS-stimulated PBMC isolated from patients at baseline, at 48 h (Early), and after treatment (End) with intravenous infusion (2 mL/kg) of either placebo (black symbols NaCl; n=12) or n-3 PUFA emulsion containing 10 g of fish oil per 100 mL (blue symbols, n=10). Results are expressed as mean ± SEM. Statistical analyses were performed with 2-way ANOVA for repeated measures and post hoc testing. Statistical significance was set at P=0.05. * p<0.05; **p<0.01compared to baseline and ^#^p<0.05 between placebo and n-3 PUFA treatment.

#### Inflammatory activation of PBMC

The release of pro-inflammatory cytokines from PBMCs isolated from the trial patients was low under basal conditions measured in supernatants from unstimulated PBMC and was significantly increased with LPS stimulation *ex vivo* (Fig 4). The *ex vivo* LPS-stimulated release of cytokines that have been most strongly associated with COVID-19 in previous reports, e.g. IL1-β, IL-6, IL-12p70 and TNF-α, was not significantly different between PBMC derived from the n-3 PUFA and placebo groups (Fig 4). In contrast, the LPS-stimulated cytokine release from PBMC was blunted in cortisone-treated patients without apparent alterations by n-3 PUFA in either the absence or presence of cortisone treatment (Fig 4).

**Fig 4.**
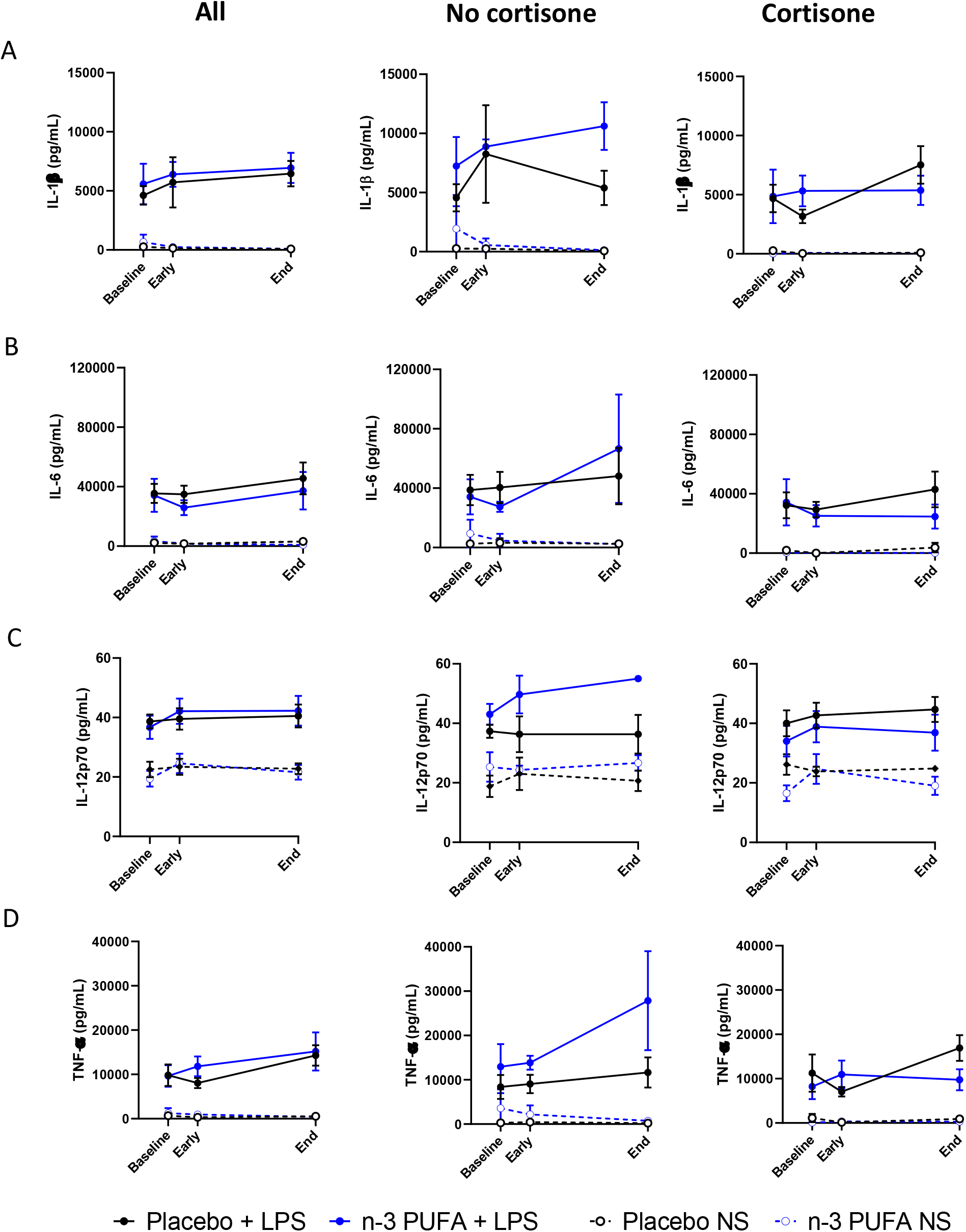
Pro-inflammatory cytokines (A) IL-1b, (B) IL-6, (C) IL-12p70 and (D) TNF-a in supernatants from non-stimulated (NS, dotted lines) and, LPS-stimulated (LPS, solid lines) PBMC isolated from patients at baseline, at 48 h (Early), and after treatment (End) with intravenous infusion (2 mL/kg) of either placebo (black symbols NaCl); n=12) or n-3 PUFA emulsion containing 10 g of fish oil per 100 mL (blue symbols, LPS n=10; NS n=9) for 5 days. Subgroups without (middle panel; placebo n=6 and n-3 PUFA n=3) or with (right panel; placebo n=6, n-3 PUFA+LPS n=7 and n-3 PUFA NS n=6) concomitant cortisone treatment are shown in middle and right panels, respectively. Results are expressed as mean ± SEM. Statistical analyses were performed with 2-way ANOVA for repeated measures.

#### Interferon-responsive gene transcription in WISH cells after treatment with patient serum

In COVID-19, increased cytokine release is accompanied by decreased interferon release (17). The release of the type 1 interferons (IFN-1) α and β from PBMCs derived from COVID-19 patients in the present trial were low at baseline with a small but significant increase after LPS stimulation (Supplementary Fig 3). The type II interferon IFN-γ was secreted at higher levels and significantly increased after LPS stimulation. There were no significant differences between the treatment groups (Supplementary Fig 3). To increase the sensitivity and to assess the functional consequences of IFN signaling, the specific IFN-responsive WISH cell line was used to determine IFN-1 responses in n-3 PUFA- and placebo-treated patients. Stimulating WISH cells with serum from trial patients revealed a time-dependent differential transcription of the IFN-1 response gene MX1 between the groups (Fig 5A). While the IFN-1-response was preserved after stimulation with serum from n-PUFA treated patients, a time-dependent decrease was observed in cells stimulated with placebo serum (Fig 5A, left panel). To exclude that the observed cellular effects were due to a direct effect of the higher n-3 PUFA concentrations in the active treatment compared with the placebo group (Fig 2), the serum-stimulation experiments were repeated with WISH cells that were grown for three consecutive passages in the presence of n-3 PUFA (MEM supplemented with 0.01% n-PUFA emulsion). These experiments revealed similar differences between the groups (Fig 5A, middle panel), excluding that exogenous n-3 PUFA stimulation affected the IFN-1 response genes and supporting that the observed effects reflected the systemic effects on IFN-1 by n-3 PUFA treatment of COVID-19. A subgroup analysis revealed that the lowest expression of IFN-1-response gene transcription was observed for cortisone-treated patients in the placebo group (Fig 5A, right panel). In contrast, IFN-1 responses were preserved in WISH cells treated with serum from n-3 PUFA-treated patients regardless of concomitant cortisone-treatment or not (Fig 5A, right panel).

**Fig 5.**
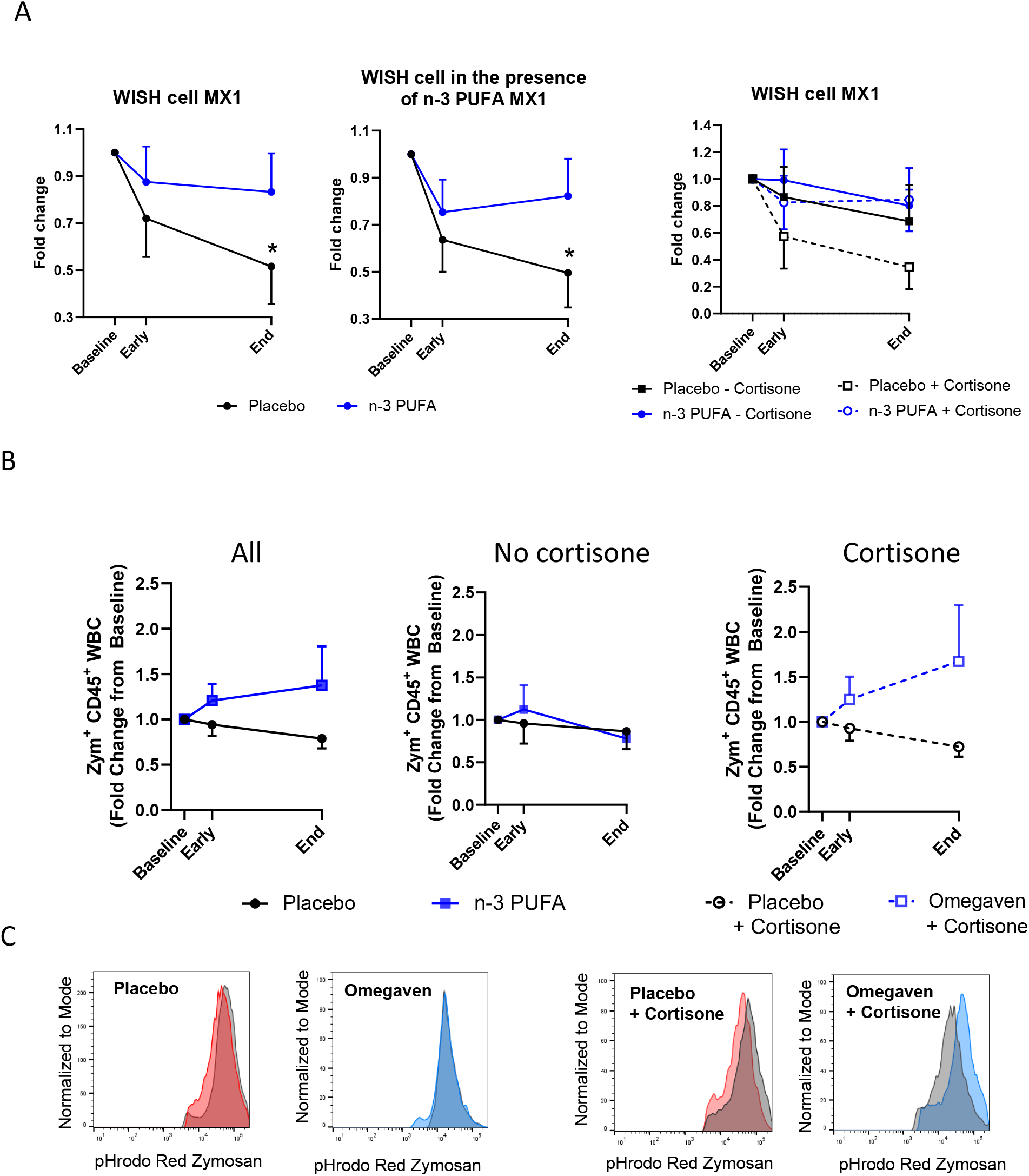
**(A)** RT-qPCR analysis of MX1 mRNA expression in WISH cells stimulated with patient serum in the absence (right panel) or presence (middle panel) of exogenous n-3 PUFA (placebo n=12; n-3 PUFA n=10). Subgroups with (dotted line; placebo n=6, n-3 PUFA n=7) and without (solid lines; placebo n=6, n-3 PUFA n=3) concomitant cortisone treatment are shown in right panel. **(B)** Fold change in phagocytosis of pHrodo-red labeled zymosan by human peripheral blood leukocytes (CD45^+^) after incubation for 45 min at 37°C (placebo n=11; n-3 PUFA n=9). Subgroups without (middle panel; placebo n=5 and n-3 PUFA n=3) or with (right panel; placebo n=6, n-3 PUFA n=6) concomitant cortisone treatment are shown in middle and right panels, respectively. (C) Representative histograms from each group and subgroup at End. Serum and whole blood were derived from patients at baseline, at 48 h (Early) and after treatment (End) with intravenous infusion (2 mL/kg) of either placebo (black symbols NaCl) or n-3 PUFA emulsion containing 10 g of fish oil per 100 mL (blue symbols). Subgroups with (dotted line) and without concomitant cortisone treatment are shown in A, right panel and B middle panel.

#### Phagocytosis after n-3 PUFA treatment

Phagocytosis is one of the key events to permit an adequate resolution of inflammation. Therefore, treatment effects on phagocytic capacity were evaluated by *ex vivo* stimulation and flow cytometry analysis of blood leukocytes derived from COVID-19 patients in the present trial. Blood leukocytes derived from n-3 PUFA-treated patients exhibited a trend towards enhanced phagocytosis of Zymosan particles, which was driven by increased phagocytosis in the group with a combined cortisone and n-3 PUFA treatment (Fig 5B).

### Descriptive endpoint for clinical course of disease

Length of hospital stay is shown in Supplemental Fig 4.

## Discussion

This single-blind randomized controlled proof-of-concept trial of i.v. n-3 PUFA in COVID-19 met its primary endpoint of changes in inflammatory biomarkers for white blood cell counts and lipid metabolites, whereas observed changes in CRP and cytokines were non-significant. We present here also the results of the exploratory analysis of the underlying mechanisms using multiple *in vitro* experimental approaches on the clinically obtained samples. The immunothrombosis interaction was significantly reduced by n-3 PUFA compared with placebo. In addition, n-3 PUFA treatment preserved IFN-1 signaling and phagocytosis. Taken together, these results point to a beneficial immunomodulation by n-3 PUFA in COVID-19 with implications for systemic inflammation, immunothrombosis, and counteraction of cortisone-induced immunosuppression.

The present trial included multimorbid older subjects, which are at a particular high risk of adverse outcomes in COVID-19 and may not always benefit from advanced treatments. The results show beneficial effects of n-3 PUFA on NLR, which is a marker of poor prognosis in COVID-19 (18). This observation was driven by a significantly increased lymphocyte count after n-3 PUFA treatment compared with placebo. The study was not adequately powered to detect effects on clinical course of disease and therefore length of hospital stay is shown in a descriptive analysis. Finally, i.v. n-3 PUFA administration was safe and feasible during hospitalization for COVID-19.

This is the first trial of n-PUFA in COVID-19 that reports the changes in the PUFA metabolome as assessed by Liquid chromatography–mass spectrometry (LC-MS/MS) -based lipid metabolite quantification. The bioavailability of i.v. n-3 PUFA administration in COVID-19 was confirmed by increased EPA and DHA plasma levels. The n-3 PUFA metabolome was in addition robustly increased in the plasma, predominantly manifested for EPA-derived metabolites. The identification of 18-HEPE among the critical PUFA metabolites that change as a result of the n-3 PUFA treatment of COVID-19 supports previous experimental studies of n-3 PUFA supplementation (19, 20). 18-HEPE is the precursor for the short-lived, locally produced, specialized proresolving mediators (SPM) resolvins (Rv) E1 (RvE1) (21) and E3 (22), which may contribute to control inflammation in COVID-19 (23, 24). Few clinical studies have however previously addressed changes in SPM and their precursors after n-3 PUFA supplementation in healthy volunteers with contradictory results (25, 26), and for limited number of subjects from cardiovascular n-3 PUFA trials (27).

In addition to the increased SPM precursor, the present trial identified changes in PUFA metabolites derived from double bond epoxidation followed by conversion to the corresponding vicinal diols by the enzyme soluble epoxide hydrolase (sEH). Treatment with n-3 PUFA increased the circulatory levels of the EPA-derived diols 11,12- and 14,15-diHETE, while the corresponding LA-derived diols of leukotoxin 9,10-diHOME and isoleukotoxin 12,13-diHOME decreased. Leukotoxins and their diols have been identified as key effectors in adult respiratory distress syndrome (ARDS) (28, 29) and detected at increased levels in COVID-19 patients with severe pulmonary involvement (30). The results of the present study extend the therapeutic implications of n-3 PUFA treatment to substrate competition for sEH-catalyzed formation of leukotoxins to support the emerging notion of sEH as a COVID-19 therapeutic target (2). Beneficial effects of decreased leukotoxin diols by n-3 PUFA treatment are supported by the increased lymphocyte counts in n-3 PUFA treated COVID-19 patients in the present trial.

Leukocyte–platelet aggregates are of importance for the severity of ARDS (31) and have emerged as of importance for pathophysiology and thrombotic complications of COVID-19 (32) through an immunothrombosis interaction (33, 34). In support of representing an additional targeted pathway in the immunomodulation by n-3 PUFA, the present trial revealed decreased platelet leukocyte aggregates in the n-PUFA treatment group, which was driven by reduced formation of neutrophil platelet aggregates. Furthermore, PMBCs isolated during and after n-3 PUFA treatment released significantly lower levels of PDGF and RANTES, which are potent mediators of leukocyte-platelet aggregates (35). Taken together, these results point to improved immunothrombosis in COVID-19 by n-3 PUFA.

The subjects included in this trial required hospitalization but did not exhibit signs of ongoing cytokine storm given the moderate cytokine increase in plasma and low spontaneous release from isolated PBMCs. Nevertheless, when mimicking the cytokine release syndrome by *ex vivo* LPS stimulation, cytokine release was significantly increased, without significant attenuation in n-3 PUFA treated subjects. These findings contrast with sepsis patients, in which PBMCs isolated after intravenous n-3 PUFA treatment reduce the LPS-stimulated cytokine release by approximately 30% (11). The present study was performed during the introduction of cortisone treatment (5), which was equally distributed within the placebo and active treatment groups. Lower stimulated cytokine release was observed in PBMCs isolated from cortisone-treated COVID-19 patients, confirming the anti-inflammatory effects of cortisone treatment on the release of the mediators identified as key in the cytokine release syndrome (6).

Concerns have been raised of cortisone-induced anti-inflammation also being indicative of an immunosuppression (36). One important beneficial immune response to consider for an optimal resolution of COVID-19 inflammation is the anti-viral effects of IFN-1. Particularly the enhanced inflammation and more severe disease reported in older patients with comorbidities has been linked to suppressed IFN-1 (37). The latter observation was extended in the present study to a low IFN-1 release from isolated PBMCs in older patients. Through a translational approach examining MX1 expression after stimulation of specific IFN-responsive WISH cell with serum from the trial participants, we show that IFN-1 responses were retained in patients treated with n-3 PUFA whereas a time-dependent decrease in the transcriptional IFN-1 response was observed in cells treated with placebo serum. Importantly, cortisone treatment suppressed IFN-1 responses to a higher degree in placebo-compared with n-3 PUFA treated patients. These results provide an initial suggestion for a retained IFN-1 response with concomitant n-3 PUFA administration with cortisone-treatment of COVID-19

In further support of n-3 PUFA as a promoter of the resolution of COVID-19-induced inflammation, the decreased phagocyte function in COVID-19 is restored *in vitro* after exogenous administration of n-3 PUFA-derived proresolving lipid mediators (38). Interestingly, the present study extends those *in vitro* effects to a clinically relevant effect of i.v. n-3 PUFA administration in COVID-19 by showing that zymosan phagocytosis was enhanced in whole blood from n-3 PUFA compared with placebo-treated patients. The apparent most prominent phagocytosis increase was observed in the group with a combined cortisone and n-3 PUFA treatment, further supporting a beneficial synergetic effect of cortisone and n-3 PUFA treatment in COVID-19. The stimulated phagocytosis by n-PUFA suggest retaining of crucial immune functions for clearance mechanisms to sustain the resolution response and counteract against cortisone-induced immunosuppression.

The main limitation is the low number of participants. It is important to consider that the study was a proof-of-concept trial powered to detect significant effects on leukocyte, lipid, and protein inflammatory biomarkers and designed to allow mechanistic exploration of n-3 PUFA metabolism and cellular effects (12). Larger studies are nevertheless needed to determine if the significantly improved NLR by n-3 PUFA translates into better clinical outcomes in COVID-19. The clinical outcomes were not statistically tested in the present trial according to the *a priori* study plan and were therefore only presented in descriptive analyses. The generalisability of identified lipid metabolites was not established in the present study. The trial was performed during the introduction of cortisone, which may have influenced the cytokine-release to prevent detecting n-3 PUFA-induced effects. The subgroups of n-3 PUFA and placebo groups with or without cortisone are small and were used only for the exploratory mechanistic experiments. The older study population with multiple comorbidities and moderate COVID-19 may limit the extrapolation of the results to younger patients and severe COVID-19 cases.

In summary, the primary outcome indicated a beneficial cellular immune response by i.v. n-3 PUFA treatment of COVID-19 detected as lowered NLR. The significantly altered the plasma PUFA metabolite profiles with increased proresolving mediator precursor levels and decreased leukotoxin-diols supported the increased lymphocyte count by n-3 PUFA treatment. The exploratory cellular experiments to decipher the molecular mechanisms identified a beneficial immune response with decreased immune-thrombosis, enhanced phagocytosis and retained IFN-1 signaling in blood and plasma derived from n-3 PUFA-compared samples from placebo-treated COVID-19 patients. These results suggest direct beneficial effects and a restriction of the cortisone-induced immunosuppression by n-3 PUFA in COVID-19. In total, this proof-of-concept trial points to possible additive beneficial effects of n-3 PUFA treatment in COVID-19 on top of current treatment recommendations, which warrant further exploration in larger trials. In particular for the vulnerable older COVID-19 patients with multiple comorbidities, which tolerated and responded to n-3 PUFA treatment.

## Data Availability

Individual participant data that underlie the results reported will be shared, after deidentification, with researchers who provide a methodologically sound proposal.
Time Frame: Beginning 9 months following article publication and finishing 36 months following article publication.
Access Criteria: Investigators interested in data should contact the corresponding author.

## Conflicts of interest

None declared

## Sources of Funding

The study received a research grant from King Gustaf V and Queen Victoria Freemason Foundation. The investigators were supported by the Swedish Research Council (Grant number 2019-01486) and the Swedish Heart and Lung Foundation (grant numbers 20180571, 20190625; 20190196; 20200693; 20210519). The sources of funding had no access to the study data and no role in the design, implementation, or reporting.

---

**Supplemental Figure 1:**
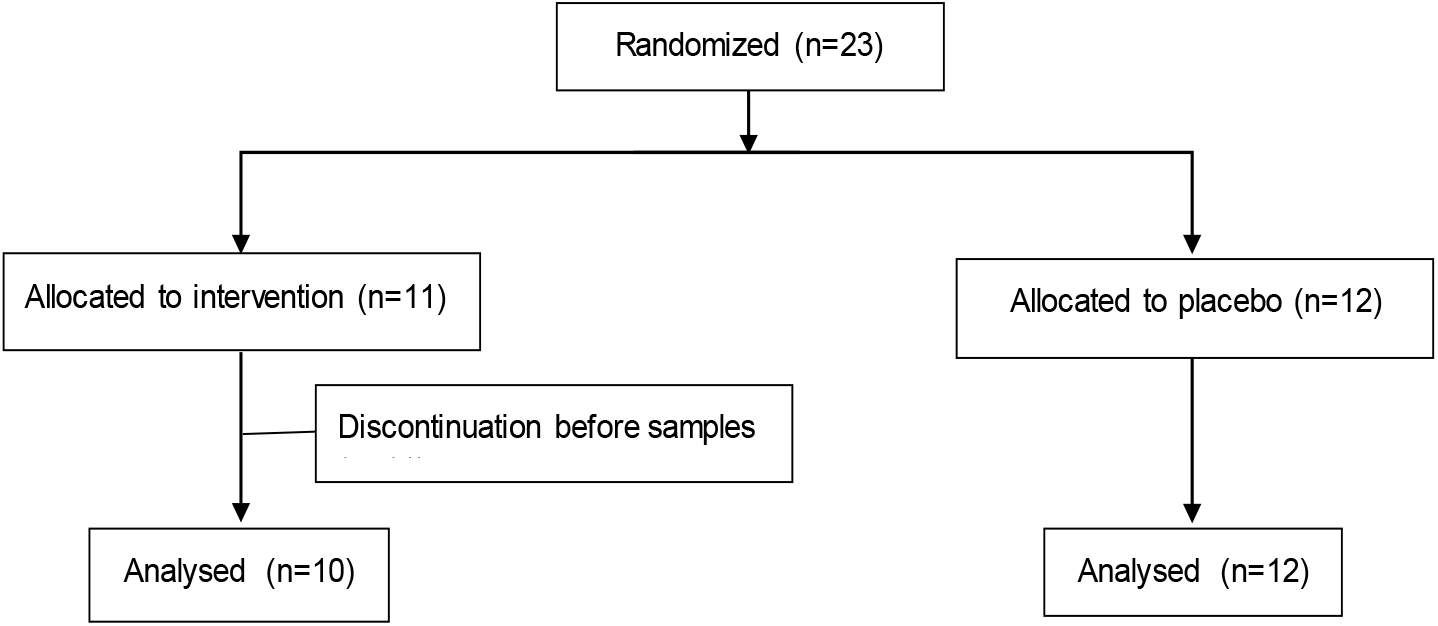

**Supplemental Figure 2:**
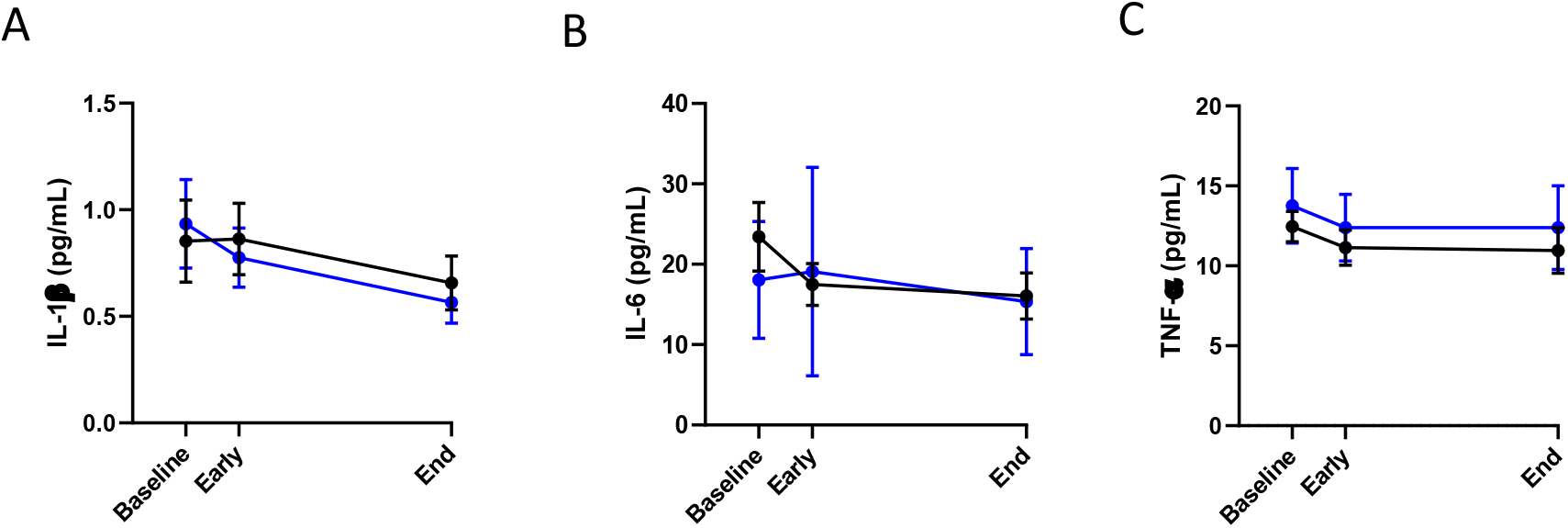
(A) IL-1β, (B) IL-6 and (C) TNF-α levels (pg/mL) in patient plasma. Cytokines were measured in plasma collected from patients at baseline, 48 h (Early) and after treatment (End) with intravenous infusion (2 mL/kg) of either placebo (NaCl; black n=12) or n-3 PUFA emulsion containing 10 g of fish oil per 100 mL (blue n=10). Results are expressed as mean ± SEM. Statistical analyses were performed with 2-way ANOVA for repeated measures and statistical significance was set at P=0.05.

**Supplemental Figure 3.**
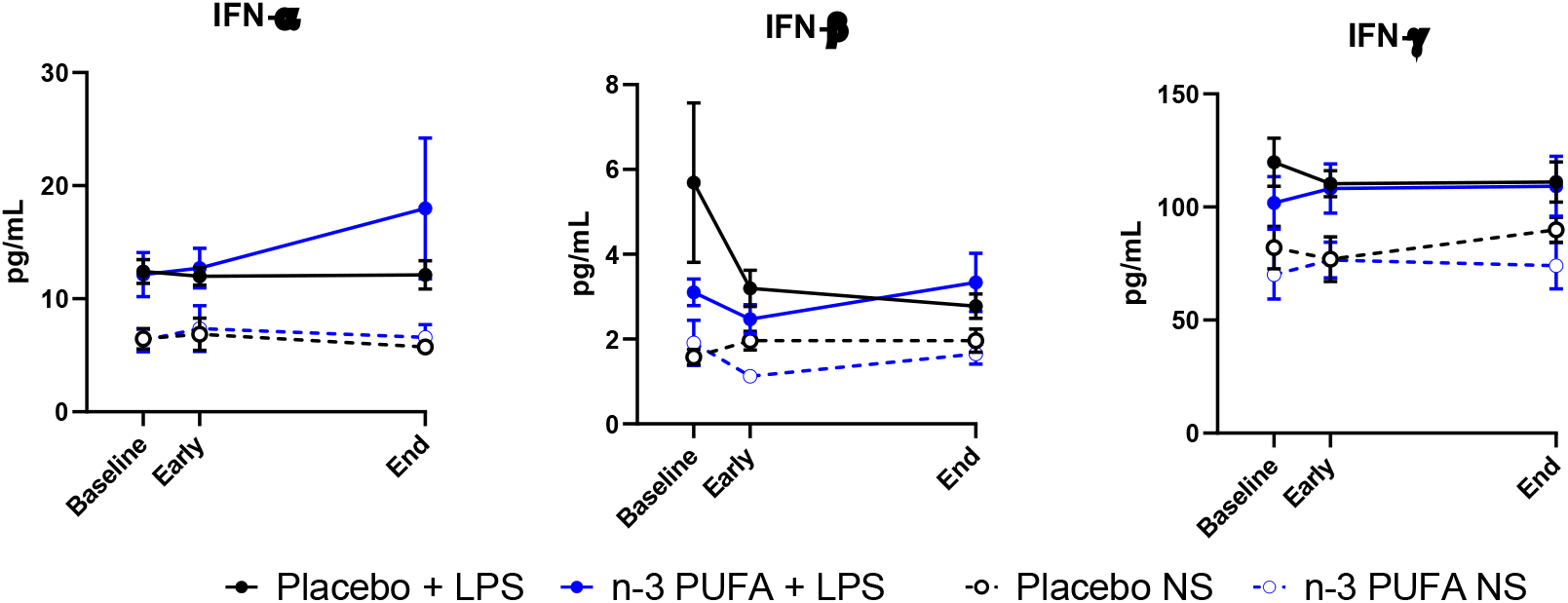
Interferon (IFN) α, β, and γ in supernatants from non-stimulated (NS, dotted lines) and, LPS-stimulated (LPS, solid lines) PBMC isolated from patients at baseline, at 48 h (Early), and after treatment (End) with intravenous infusion (2 mL/kg) of either placebo (black symbols NaCl); n=12) or n-3 PUFA emulsion containing 10 g of fish oil per 100 mL (blue symbols, LPS n=10; NS n=9) for 5 days. Results are expressed as mean ± SEM.

**Supplemental Figure 4:**
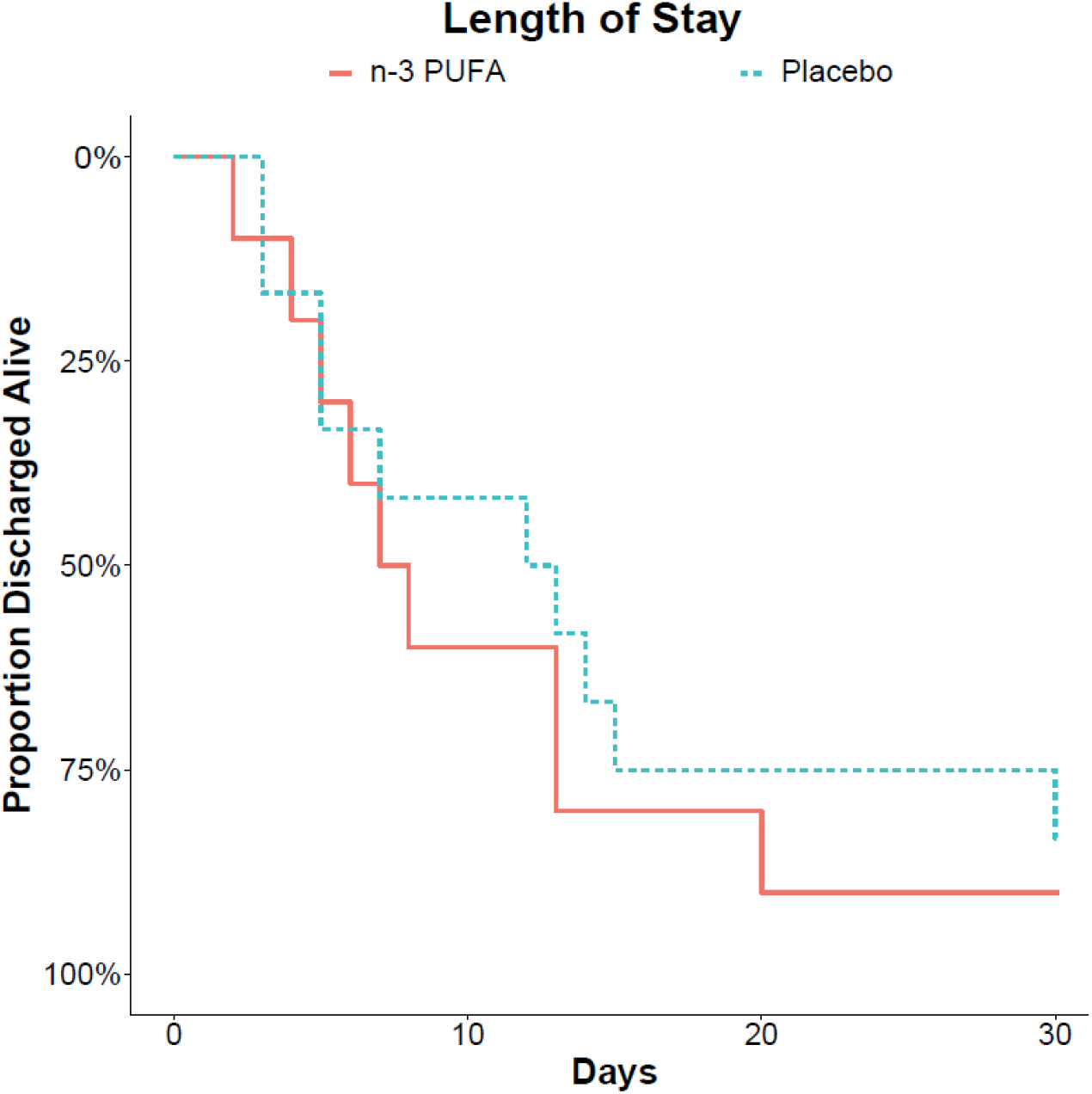
Length of hospital stay after start of treatment with intravenous infusion (2 mL/kg) of either placebo (NaCl; dotted blue line n=12) or n-3 PUFA emulsion containing 10 g of fish oil per 100 mL (solid red line n=10).

**Supplementary Table 1:** Mass spectrometry parameters for the additional compounds added to the eicosanoid method reported in *Anal Chem* 2018; 90: 10239-10248

| *Compound* | *Parent (M-1; m/z)* | *Product (M-1; m/z)* | *CE (eV)* | *Cone V* |
| --- | --- | --- | --- | --- |
| *16,17-EpDPA* | 343.0 | 188.8 | 14 | 30 |
| *7,8-EpDPA* | 343.0 | 188.8 | 12 | 30 |
| *14,15-EpETE* | 317.0 | 174.7 | 12 | 30 |
| *16-17-DiHDPA* | 361.0 | 188.8 | 18 | 30 |
| *7,8-DiHDPA* | 361.0 | 188.8 | 16 | 30 |
| *8,9-EpETE* | 317.0 | 160.7 | 12 | 30 |
| *11,12-EpETE* | 317.0 | 166.7 | 12 | 30 |
| *10,11-EpDPA* | 343.0 | 152.6 | 12 | 30 |
| *13,14-EpDPA* | 343.0 | 152.6 | 16 | 30 |
| *11,12-DiHETE* | 335.0 | 166.7 | 14 | 30 |
| *10-11-DiHDPA* | 361.0 | 182.7 | 16 | 30 |
| *13-14-DiHDPA* | 361.0 | 188.8 | 16 | 30 |

**Supplementary Table 2.** Lipid mediator metabolites from the n-6 PUFA AA in patient plasma. Data is expressed as mean (SEM) in pg/mL of plasma. P-values were derived from the mixed effects ANOVA containing intervention (Interv; n-PUFA vs. placebo) and progress (early and end samples) as a within-subjects effect and as a between subjects effect. Intervention:progress (Interv:Prog) represents the significance of the interaction between these two terms. *Abbreviations: HETE, Hydroxyeicosatetraenoic acid; KETE, Oxo-eicosatetraenoic acid; HHTrE, Hydroxyheptadecatrienoic acid; DiHETE, Dihydroxyeicosatetraenoic acid; EpETrE, Epoxyeicosatrienoic acid; DiHETrE, Dihydroxyeicoatrienoic acid*

|  | n-PUFA (n=9) |  |  |  | Placebo (n=12) |  |  |  | P-values |  |  |
| --- | --- | --- | --- | --- | --- | --- | --- | --- | --- | --- | --- |
|  | Baseline | Early | End |  | Baseline | Early | End |  | Interv | Progress | Interv:Progr |
| **AA Metabolome** |  |  |  |  |  |  |  |  |  |  |  |
| 20-HETE | 7.35 (1.9) | 10.13 (1.7) | 9.48 (1.49) |  | 10 (2.38) | 11.55 (2.06) | 10.6 (2.55) |  | 0.641 | 0.582 | 0.91 |
| 19-HETE | 7.24 (0.99) | 8 (1.44) | 7.54 (0.8) |  | 7.05 (0.92) | 7.92 (1) | 6.58 (0.82) |  | 0.688 | 0.207 | 0.534 |
| 15-HETE | 11.93 (2.24) | 10.43 (1.8) | 7.9 (0.73) |  | 7.22 (0.95) | 8.81 (1.28) | 9.4 (1.46) |  | 0.971 | 0.243 | 0.068 |
| 12-HETE | 352.2 (79.2) | 225.4 (46.8) | 309 (71.4) |  | 257 (51.0) | 329 (80.6) | 313 (59.3) |  | 0.525 | 0.511 | 0.332 |
| 11-HETE | 3.23 (0.28) | 3.1 (0.43) | 3.42 (0.66) |  | 2.84 (0.54) | 3.33 (0.71) | 3.36 (0.7) |  | 0.923 | 0.445 | 0.522 |
| 9-HETE | 1.04 (0.51) | 1.01 (0.51) | 1.99 (0.47) |  | 0.89 (0.37) | 1.58 (0.4) | 1 (0.36) |  | 0.704 | 0.487 | 0.012 |
| 8-HETE | 4.76 (0.62) | 4.6 (0.75) | 4.87 (0.68) |  | 3.61 (0.39) | 4.29 (0.4) | 4.13 (0.27) |  | 0.437 | 0.865 | 0.494 |
| 5-HETE | 33.28 (5.77) | 35.73 (9.05) | 23.96 (4.58) |  | 18.5 (1.93) | 22.68 (2.83) | 19.2 (1.93) |  | 0.119 | 0.072 | 0.314 |
| 15-KETE | 1.2 (0.16) | 1.06 (0.16) | 0.89 (0.16) |  | 0.78 (0.16) | 0.81 (0.18) | 0.84 (0.15) |  | 0.5 | 0.465 | 0.284 |
| 12-KETE | 1.23 (0.27) | 0.8 (0.16) | 0.65 (0.2) |  | 0.97 (0.27) | 1.36 (0.28) | 0.92 (0.18) |  | 0.146 | 0.08 | 0.367 |
| 5-KETE | 1.16 (0.3) | 1.4 (0.44) | 1.14 (0.23) |  | 1.11 (0.1) | 0.79 (0.19) | 1.1 (0.18) |  | 0.326 | 0.903 | 0.133 |
| PGE<sub>2</sub> | 3.73 (0.98) | 3.39 (0.83) | 5.95 (1.63) |  | 2.92 (1.26) | 4.38 (1.95) | 4.95 (1.67) |  | 0.997 | 0.063 | 0.224 |
| 13,14-dihydro-15-keto-PGE<sub>2</sub> | 0.14 (0.06) | 0.19 (0.07) | 0.12 (0.05) |  | 0.11 (0.03) | 0.15 (0.06) | 0.14 (0.05) |  | 0.854 | 0.425 | 0.486 |
| 8-isoPGE<sub>2</sub> | 0.3 (0.06) | 0.5 (0.31) | 0.21 (0.11) |  | 0.18 (0.06) | 0.36 (0.2) | 0.19 (0.07) |  | 0.734 | 0.1 | 0.668 |
| PGD<sub>2</sub> | 1.01 (0.37) | 0.81 (0.33) | 2.36 (0.6) |  | 1.01 (0.73) | 1.56 (0.75) | 1.9 (0.76) |  | 0.879 | 0.003 | 0.041 |
| PGF<sub>2α</sub> | 7.06 (1.3) | 6.07 (1.21) | 10.65 (2.62) |  | 4.63 (1.73) | 6.76 (2.49) | 7.14 (1.89) |  | 0.626 | 0.061 | 0.109 |
| PGB<sub>2</sub> | 0.37 (0.07) | 0.42 (0.12) | 0.53 (0.16) |  | 0.36 (0.09) | 0.31 (0.07) | 0.39 (0.07) |  | 0.384 | 0.073 | 0.762 |
| 12-HHTre | 14.0 (5.49) | 18.8 (8.81) | 21.5 (8.51) |  | 20.0 (13.3) | 22.0 (14.2) | 25.7 (13.8) |  | 0.831 | 0.558 | 0.917 |
| TXB<sub>2</sub> | 12.3 (5.33) | 18.81 (9.2) | 19.27 (8.35) |  | 11.6 (6.27) | 15.79 (9.65) | 23.4 (10.2) |  | 0.966 | 0.483 | 0.534 |
| 11-keto-TXB<sub>2</sub> | 0.08 (0) | 0.08 (0) | 0.08 (0) |  | 0.09 (0.01) | 0.09 (0.02) | 0.09 (0.01) |  | 0.227 | 0.823 | 0.823 |
| LTE<sub>4</sub> | 2.09 (0.94) | 2.98 (2.32) | 1.07 (0.33) |  | 0.82 (0.17) | 1.4 (0.54) | 1.11 (0.51) |  | 0.533 | 0.266 | 0.414 |
| LTD<sub>4</sub> | 0.36 (0.13) | 0.29 (0.19) | 0.14 (0.08) |  | 0.17 (0.07) | 0.32 (0.14) | 0.21 (0.11) |  | 0.784 | 0.218 | 0.869 |
| LTC<sub>4</sub> | 2.67 (0.75) | 2.62 (0.74) | 3.01 (0.68) |  | 3.74 (0.47) | 4.02 (0.79) | 2.3 (0.73) |  | 0.684 | 0.346 | 0.141 |
| LTB<sub>4</sub> | 4.79 (1.34) | 6.1 (3.52) | 2.42 (0.78) |  | 1.96 (0.31) | 2.25 (0.59) | 1.87 (0.44) |  | 0.216 | 0.186 | 0.277 |
| 6-trans-LTB<sub>4</sub> | 0.47 (0.14) | 0.43 (0.21) | 0.14 (0.06) |  | 0.11 (0.03) | 0.13 (0.04) | 0.1 (0.03) |  | 0.13 | 0.099 | 0.18 |

**Supplementary Table 2. Lipid mediator metabolites from the n-6 PUFA AA in patient plasma.**
|  | n-PUFA (n=9) |  |  |  | Placebo (n=12) |  |  |  | P-values |  |  |
| --- | --- | --- | --- | --- | --- | --- | --- | --- | --- | --- | --- |
| *20-COOH-LTB<sub>4</sub>* | 4.7 (1.92) | 4.78 (2.35) | 1.47 (0.58) |  | 0.91 (0.39) | 1.56 (0.59) | 1.42 (0.56) |  | 0.183 | 0.135 | 0.167 |
| *20-OH-LTB<sub>4</sub>* | 1.33 (0.5) | 1.03 (0.59) | 0.37 (0.11) |  | 0.61 (0.28) | 0.57 (0.21) | 0.35 (0.14) |  | 0.478 | 0.103 | 0.406 |
| *5,15-DiHETE* | 0.09 (0.05) | 0.1 (0.06) | 0.02 (0) |  | 0.05 (0.03) | 0.07 (0.03) | 0.11 (0.06) |  | 0.587 | 0.613 | 0.143 |
| *14(15)-EpETrE* | 0.62 (0.25) | 0.64 (0.29) | 0.58 (0.24) |  | 0.74 (0.2) | 0.77 (0.21) | 0.59 (0.22) |  | 0.813 | 0.496 | 0.727 |
| *11(12)-EpETrE* | 0.31 (0.09) | 0.36 (0.1) | 0.26 (0.08) |  | 0.33 (0.1) | 0.25 (0.07) | 0.19 (0.09) |  | 0.382 | 0.265 | 0.77 |
| *8(9)-EpETrE* | 0.61 (0.22) | 0.6 (0.19) | 0.48 (0.17) |  | 0.52 (0.15) | 0.32 (0.14) | 0.43 (0.13) |  | 0.421 | 0.946 | 0.197 |
| *5(6)-EpETrE* | 0.53 (0.21) | 1 (0.34) | 0.63 (0.26) |  | 0.89 (0.3) | 0.49 (0.18) | 0.69 (0.21) |  | 0.475 | 0.614 | 0.103 |
| *14,15-DiHETrE* | 4.45 (0.33) | 4.55 (0.57) | 4.23 (0.37) |  | 4.41 (0.46) | 4.23 (0.37) | 3.91 (0.2) |  | 0.494 | 0.249 | 0.995 |
| *11,12-DiHETrE* | 3.77 (0.21) | 4.03 (0.36) | 3.6 (0.27) |  | 4 (0.47) | 3.85 (0.4) | 3.61 (0.29) |  | 0.834 | 0.219 | 0.721 |
| *8,9-DiHETrE* | 1.59 (0.17) | 1.57 (0.18) | 1.51 (0.15) |  | 1.48 (0.16) | 1.67 (0.21) | 1.55 (0.09) |  | 0.703 | 0.514 | 0.832 |
| *5,6-DiHETrE* | 3.43 (0.75) | 4.12 (0.95) | 3.91 (0.87) |  | 2.52 (0.39) | 3.28 (0.38) | 4.03 (0.66) |  | 0.69 | 0.565 | 0.312 |

**Supplementary Table 3.** Lipid mediator metabolites from the n-6 PUFA LA in patient plasma. Data is expressed as mean (SEM) in pg/mL of plasma. P-values were derived from the mixed effects ANOVA containing intervention (Interv; n-PUFA vs. placebo) and progress (early and end samples) as a within-subjects effect and as a between subjects effect. Intervention:progress (Interv:Prog) represents the significance of the interaction between these two terms.

|  | n-PUFA (n=9) |  |  |  | Placebo (n=12) |  |  |  | P-values |  |  |
| --- | --- | --- | --- | --- | --- | --- | --- | --- | --- | --- | --- |
|  | Baseline | Early | End |  | Baseline | Early | End |  | Interv | Progress | Interv:Progr |
| ***C18 LA & GLA Metabolomes*** |  |  |  |  |  |  |  |  |  |  |  |
| 9-HODE | 74.4±10.9 | 95.5±20.4 | 60.3±8.95 |  | 81.9±13.3 | 77.1±17.2 | 72.5±8.15 |  | 0.858 | 0.099 | 0.199 |
| 13-HODE | 100±10.6 | 112±18.2 | 83.0±12.9 |  | 116±18.4 | 103±21.0 | 104±11.1 |  | 0.776 | 0.256 | 0.212 |
| 9-KODE | 11.5±2.16 | 11±2.7 | 10.1±3.65 |  | 16.8±3.54 | 15.4±3.95 | 16.0±3.69 |  | 0.287 | 0.952 | 0.757 |
| 13-KODE | 27.2±4.65 | 24.3±4.0 | 18.5±2.82 |  | 26.6±3.39 | 25.8±3.63 | 28.5±3.73 |  | 0.218 | 0.565 | 0.126 |
| 9(10)-EpOME | 12.7±1.37 | 13.0±2.19 | 10.4±1.81 |  | 17.2±2.79 | 14.0±1.98 | 14.9±1.79 |  | 0.222 | 0.628 | 0.334 |
| 12(13)-EpOME | 26.0±2.95 | 32.4±8.55 | 20.7±3.75 |  | 33.0±5.20 | 27.0±5.24 | 29.8±4.55 |  | 0.767 | 0.404 | 0.183 |
| 9,10-DiHOME | 43.1±11.0 | 59.3±17.0 | 31.6±8.24 |  | 48.2±10.5 | 52.0±14.3 | 64.9±9.44 |  | 0.36 | 0.534 | 0.099 |
| 12,13-DiHOME | 33.1±5.25 | 41.8±10.9 | 25.0±4.39 |  | 32.5±5.94 | 30.8±6.8 | 34.0±7.99 |  | 0.904 | 0.36 | 0.187 |
| 9,10,13-TriHOME | 3.29±0.33 | 4.62±1.05 | 3.11±0.35 |  | 2.81±0.37 | 2.76±0.26 | 3.33±0.35 |  | 0.2 | 0.309 | 0.031 |
| 9,12,13-TriHOME | 5.96±2.55 | 9.50±6.07 | 1.65±0.37 |  | 3.40±1.38 | 1.70±0.36 | 9.69±8.29 |  | 0.982 | 0.991 | 0.169 |
| EKODE | 151±31.8 | 104±25.1 | 123±36.1 |  | 160±40.1 | 137±34.3 | 132±40.0 |  | 0.675 | 0.636 | 0.415 |
| 13-HOTrEy | 1.35±0.35 | 0.9±0.17 | 0.69±0.11 |  | 0.6±0.13 | 0.74±0.18 | 0.92±0.28 |  | 0.901 | 0.912 | 0.145 |
| ***C20 DGLA and EDE Metabolomes*** |  |  |  |  |  |  |  |  |  |  |  |
| 15-HETrE | 1.43±0.18 | 1.09±0.12 | 0.91±0.09 |  | 1.28±0.13 | 1.68±0.27 | 1.61±0.31 |  | 0.065 | 0.325 | 0.655 |
| 8-HETrE | 1.79±0.24 | 1.67±0.31 | 1.66±0.24 |  | 1.76±0.14 | 2.14±0.18 | 1.61±0.17 |  | 0.447 | 0.108 | 0.119 |
| 5-HETrE | 2.05±0.3 | 1.88±0.48 | 1.26±0.22 |  | 1.66±0.27 | 1.79±0.19 | 1.44±0.18 |  | 0.883 | 0.054 | 0.559 |
| PGE<sub>1</sub> | 0.21±0.07 | 0.12±0.04 | 0.2±0.05 |  | 0.15±0.04 | 0.15±0.06 | 0.17±0.05 |  | 0.945 | 0.013 | 0.174 |
| PGD<sub>1</sub> | 0.02±0 | 0.02±0 | 0.02±0 |  | 0.04±0.02 | 0.06±0.02 | 0.05±0.02 |  | 0.223 | 0.4 | 0.4 |
| TXB<sub>1</sub> | 0.82±0.26 | 1.2±0.46 | 0.68±0.35 |  | 2.2±0.85 | 1.56±0.57 | 1.38±0.58 |  | 0.477 | 0.094 | 0.419 |
| 11-HEDE | 0.66±0.19 | 0.68±0.29 | 0.55±0.2 |  | 0.75±0.19 | 0.73±0.27 | 0.73±0.21 |  | 0.709 | 0.709 | 0.704 |

**Supplementary Table 3. Lipid mediator metabolites from the n-6 PUFA LA in patient plasma.**
|  | n-PUFA (n=9) |  |  |  | Placebo (n=12) |  |  | P-values |  |  |
| --- | --- | --- | --- | --- | --- | --- | --- | --- | --- | --- |
| ***C18 ALA Metabolome*** |  |  |  |  |  |  |  |  |  |  |
| *13-HOTrE* | 11.9±1.38 | 13.8±2.38 | 9.18±1.15 |  | 11.1±1.95 | 9.33±1.38 | 12.1±1.29 | 0.661 | 0.547 | 0.021 |
| *9-HOTrE* | 6.43±0.87 | 8.82±2.22 | 4.92±0.68 |  | 6.07±1.1 | 5.33±0.99 | 5.63±0.66 | 0.283 | 0.142 | 0.089 |
| *9-KOTrE* | 0.94±0.14 | 0.99±0.17 | 0.85±0.27 |  | 1.18±0.16 | 0.97±0.19 | 1.11±0.17 | 0.634 | 0.98 | 0.315 |
| *15(16)-EpODE* | 10504.9±12<br>83.89 | 16673.39±4<br>851.33 | 7657.52±13<br>67.92 |  | 10624.25±1<br>948.48 | 10771.8±42<br>78.08 | 11996.31±1<br>757.29 | 0.819 | 0.284 | 0.164 |
| *12(13)-EpODE* | 1.08±0.17 | 2.01±0.89 | 0.83±0.22 |  | 1.27±0.22 | 0.92±0.18 | 1.16±0.17 | 0.378 | 0.273 | 0.104 |
| *9(10)-EpODE* | 0.02±0 | 0.02±0 | 0.02±0 |  | 8.32±8.3 | 0.02±0 | 25.9±15.1 | 0.158 | 0.158 | 0.158 |

## Notes

### Competing Interest Statement

The authors have declared no competing interest.

### Clinical Trial

NCT04647604

### Author Declarations

Approved by approved by Swedish Ethical Review Authority (Dnr 2020-02592) and Medical Product Agency (Dnr 5.1-2020-42861).

